# Imprinted SARS-CoV-2-specific memory lymphocytes define hybrid immunity

**DOI:** 10.1101/2022.01.12.22269192

**Authors:** Lauren B. Rodda, Peter A. Morawski, Kurt B. Pruner, Mitchell L Fahning, Christian A. Howard, Nicholas Franko, Jennifer Logue, Julie Eggenberger, Caleb Stokes, Inah Golez, Malika Hale, Michael Gale, Helen Y. Chu, Daniel J. Campbell, Marion Pepper

## Abstract

Immune memory is tailored by cues that lymphocytes perceive during priming. The severe acute respiratory syndrome coronavirus 2 (SARS-CoV-2) pandemic created a situation in which nascent memory could be tracked through additional antigen exposures. Both SARS-CoV-2 infection and vaccination induce multifaceted, functional immune memory, but together they engender improved protection from disease, termed “hybrid immunity”. We therefore investigated how vaccine-induced memory is shaped by previous infection. We found that following vaccination, previously infected individuals generated more SARS-CoV-2 RBD-specific memory B cells and variant-neutralizing antibodies and a distinct population of IFN-γ and IL-10-expressing memory SARS-CoV-2 spike-specific CD4+ T cells than previously naive individuals. While additional vaccination could increase humoral memory, it did not recapitulate the distinct CD4+ T cell cytokine profile in previously naive individuals. Thus, imprinted features of SARS-CoV-2-specific memory lymphocytes define hybrid immunity.

## Introduction

The generation of immune memory is influenced by signals that B and T cells of the adaptive immune system perceive during a primary immune response. This system has evolved such that key cues from an invading pathogen or vaccine are relayed to lymphocytes so they can provide specific functional outputs to combat infection and protect from re-infection. The site of antigen encounter, specific inflammatory signals, and the number and frequency of antigen exposures all influence the resulting memory pool, yet the specific rules governing memory formation remain undefined. Understanding how the confluence of these parameters influences immune memory function and maintenance is critical for optimizing protective vaccines.

In the context of the ongoing SARS-CoV-2 pandemic, so-called “hybrid/super-immunity” induced by a combination of prior SARS-CoV-2 infection and subsequent COVID-19 vaccination, provides greater protection against re-infection and severe COVID-19 disease than either infection or vaccination alone (Abu-Raddad et al., 2021; Crotty 2021; Gazit et al., 2021; Goldberg et al., 2021; Reynolds et al., 2021). Studies investigating immune correlates of hybrid-immunity have revealed improved breadth and neutralizing ability of circulating antibodies from previously infected individuals (Cho et al., 2021; Schmidt et al., 2021; Stamatatos et al., 2021; Wang et al., 2021) but the specific changes in the cellular immune compartment associated with this immune state remain undefined. Additionally, it is not understood whether further activation of immune memory in vaccinated only individuals could achieve similar qualities, as previously infected individuals have had an additional antigen exposure.

To answer these questions, we tracked circulating SARS-CoV-2-specific antibodies and memory lymphocytes in a cohort of Naive (N) or SARS-CoV2-Previously Infected (PI) subjects over the course of three vaccinations. We focused on visualizing RBD-specific antibodies and B cells and spike (S)-specific CD4+ T cells as they are critical mediators of protection in infected individuals over a two year time period and targets of SARS-CoV-2-directed vaccines (Corbett et al., 2021; Feng et al., 2021; Gilbert et al., 2021; Khoury et al., 2021).

We find that the immune memory landscape elicited by vaccination of previously infected subjects is distinct from the immune memory of SARS-CoV-2-naive individuals. Three months after two doses of vaccination, PI individuals maintained a higher quantity of RBD-specific plasma antibodies with superior plasma neutralization of viral variants including Omicron. PI individuals also retained increased numbers of RBD-specific B cells enriched for IgG+ classical and activated MBCs compared to N individuals. However, these differences were normalized by administration of a third vaccine dose. Examination of the T cell compartment revealed that although N and PI individuals generated equivalent numbers of S-specific CD4+ T cells after vaccination, there was a profound functional skewing towards a Th1 phenotype in PI subjects. CD4+ T cell functional differences between N and PI subjects persisted following the administration of a vaccine booster. Further, although the third dose of vaccine provided a boost in circulating antibodies, we observed no increase in memory lymphocytes, indicating that the immune memory compartment is likely maximized after the two-dose regimen. Thus, our data support a model in which the priming environment induced by SARS-CoV-2 infection imprints immune memory with multiple features of enhanced type-1 antiviral immunity. These likely contribute to the increased protection associated with hybrid immunity and are not recapitulated by repeat vaccination.

## Results

### The greater humoral response to vaccination in SARS-CoV-2 previously infected compared to naive individuals is recovered by third vaccination

To investigate how prior SARS-CoV-2 infection impacts the quantity and quality of vaccine-induced SARS-CoV-2-specific immune memory, we collected plasma and peripheral blood mononuclear cells (PBMCs) from SARS-CoV-2 N and SARS-CoV-2 PI individuals before vaccination and 1 week, 3 months and 6 months after two vaccinations with BNT162b2 (Pfizer-BioNTech) or mRNA-1273 (Moderna) COVID-19 vaccines (**Figure 1A** and **Table S1)** (Baden et al., 2021; Polack et al., 2020). An average of 8 months following initial vaccination, a subset of participants received a third vaccine dose and we collected samples 2 weeks (mean 18 days, range 13-47 days) later. N participants had no detectable SARS-CoV-2 RBD-specific IgG plasma antibodies above a cutoff set by historical negative controls prior to vaccination (**Figure S1A**). PI participants reported a positive SARS-CoV-2 test and mildly symptomatic disease an average of 9 months before their pre-vaccination blood draw.

**Figure 1.**
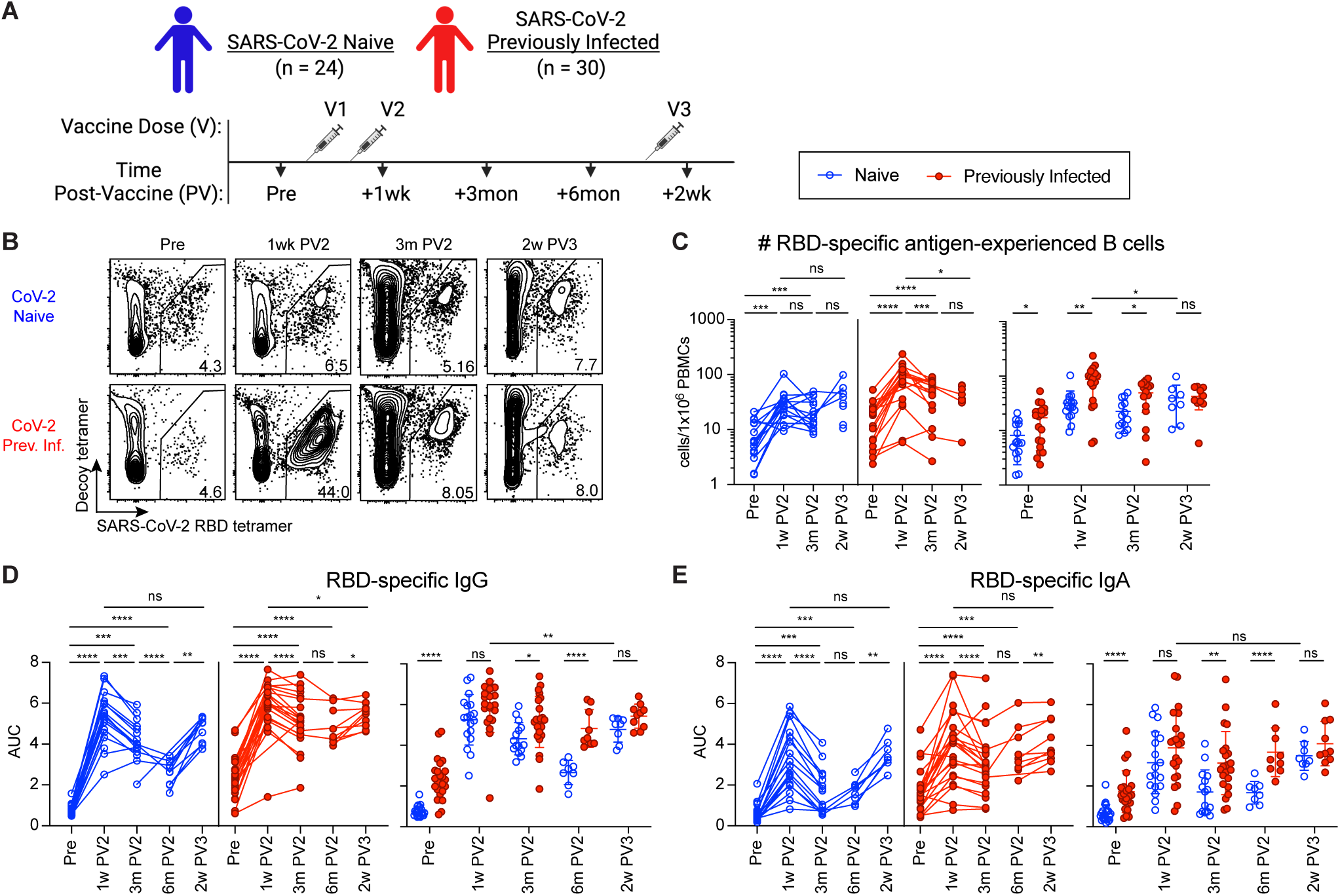
The greater humoral response to vaccination in SARS-CoV-2 previously infected compared to naive individuals is recovered by third vaccination. (A) Timeline of blood draws from SARS-CoV-2 naive (N) and previously infected (Prev. Inf., PI) analyzed in this study relative to vaccinations. (B) Representative gating on CD19^+^CD38^lo^ B cells for RBD-tetramer^+^Decoy^-^ SARS-CoV-2 RBD-specific B cells from N and PI (Prev. Inf.) PBMCs at the indicated time points pre-vaccination (Pre), 1 week post-2nd vaccination (1w PV2), 3 months post-2nd vaccination (3m PV2) and 6 months post-2nd vaccination (6m PV2). (C) Number of RBD-specific antigen experienced (ag-exp.) B cells (CD21^+^CD27^+^ and CD21^-^CD27^+/-^) in N (blue) and PI (red) PBMCs at the indicated time points. (D and E) ELISA area under the curve (AUC) for RBD-specific IgG (D) and IgA (E) plasma antibody from N and PI individuals at indicated time points. Statistics for unpaired data determined by 2-tailed Mann-Whitney tests and, for paired data, by 2-tailed Wilcoxon signed-rank tests: not significant (ns), \**P* < 0.05, \*\**P* < 0.005, \*\*\**P* < 0.0005, and \*\*\*\**P* < 0.0001. Error bars represent mean and SD. See also: Supplementary Table 1 and Figure S1.

To determine the distinct humoral features of hybrid immunity, we first compared the circulating SARS-CoV-2 RBD-specific B cells in N and PI participants pre- and post-vaccination. Tetramer enrichment allowed us to identify rare SARS-CoV-2 RBD-specific B cells from PBMCs to track the memory response (**Figure S1B**). Pre-vaccination, PI individuals had significantly more RBD-specific antigen-experienced B cells (CD21^+^CD27^+^ and CD21^-^CD27^+/-^)(Cancro and Tamayko, 2021) (**Figures 1B** and **1C, S1C**) and plasma RBD-specific IgG and IgA than N individuals (**Figures 1D** and **1E**), reflecting the sustained memory response to their previous SARS-CoV-2 infection (**Figures 1B** and **1C, S1C**) (Rodda et al., 2021, Dan et al., 2021). Two dose vaccination induced a robust humoral response in both N and PI individuals including increased numbers of RBD-specific antigen-experienced B cells (**Figure 1C**), an acute burst of RBD-specific plasmablasts (**Figure S1D**) and increased titers of RBD-specific IgG and IgA antibody (**Figures 1D** and **1E**). The numbers of RBD-specific antigen-experienced B cells and RBD-specific antibodies in both groups contracted to levels higher than those at pre-vaccination. However, PI individuals formed and retained higher numbers of RBD-specific antigen-experienced B cells and RBD-specific IgG and IgA than N individuals at 3 and 6 months post-vaccination (**Figures 1C-E**). These findings corroborate recent work and support the likely functional role of the elevated RBD-specific antibody and MBCs in hybrid immunity (Goel et al., 2021).

To determine if a third vaccine dose could overcome these differences by engaging the memory response in N individuals in additional expansion, we measured the humoral response in both groups 2 weeks after a third dose. While RBD-specific B cells in N individuals responded by increasing RBD-specific IgG and IgA antibody to the levels in PI individuals (**Figures 1D-F**), the antibody response in N individuals to a third SARS-CoV-2 antigen exposure did not match the acute response in PI individuals to their third exposure (vaccination 2) (**Figures 1C and 1D**). This muted response to a third exposure in N and PI individuals most likely reflects efficient clearance of this smaller antigen dose by the high quantity of antigen-specific memory now established in both groups. However, data from a further memory time point post-3rd vaccination is required to determine if this muted response is sufficient to induce a sustained expansion of RBD-specific MBCs and plasma antibody in N individuals to match that in PI individuals.

### Vaccination induces a robust and durable virus-specific CD4+ T cell response irrespective of prior SARS-CoV-2 infection

CD4+ T cell responses to vaccination were also assessed in N and PI individuals using an activation induced marker (AIM) assay based on expression of CD69 and CD137 to detect SARS-CoV-2 specificity (Tarke et al., 2021) (**Figures S2A-S2D**). PBMCs from N or PI donors isolated before and after vaccination were re-stimulated in the presence of peptide pools optimized for the induction of MHC class II-dependent CD4+ T cell responses (15-mers), and containing predicted T cell epitopes across a range of common HLA haplotypes against the viral membrane and nucleocapsid (M/N) or spike (S) proteins (**Table S2**).

Pre-vaccination, CD4+ T cells from PI donors showed responses to both M/N and S, consistent with prior SARS-CoV-2 infection and the presence of memory cells (**Figures 2A** and **2B**), whereas responses in N donors were undetectable above individual donor background (**Figures S3A** and **S3B**). Throughout the course of vaccination, responses to M/N remained negative in N donors, but endured, undiminished in PI donors at all time points, indicating that SARS-CoV-2-specific CD4+ T cells primed during infection, persist at least 18 months following initial antigen exposure (**Figure 2B**). Following initial two-dose vaccination, S-reactive AIM+CD4+ T cells were significantly elevated above pre-vaccination levels in both N and PI individuals, with no significant differences in peak CD4+ T cell response between groups (**Figures 2B**, **S3C**, and **S3D**). Therefore COVID-19 vaccination induces numerically equivalent populations of S-specific cells in all subjects, regardless of prior SARS-CoV-2 exposure status, that persist for at least three months.

**Figure 2.**
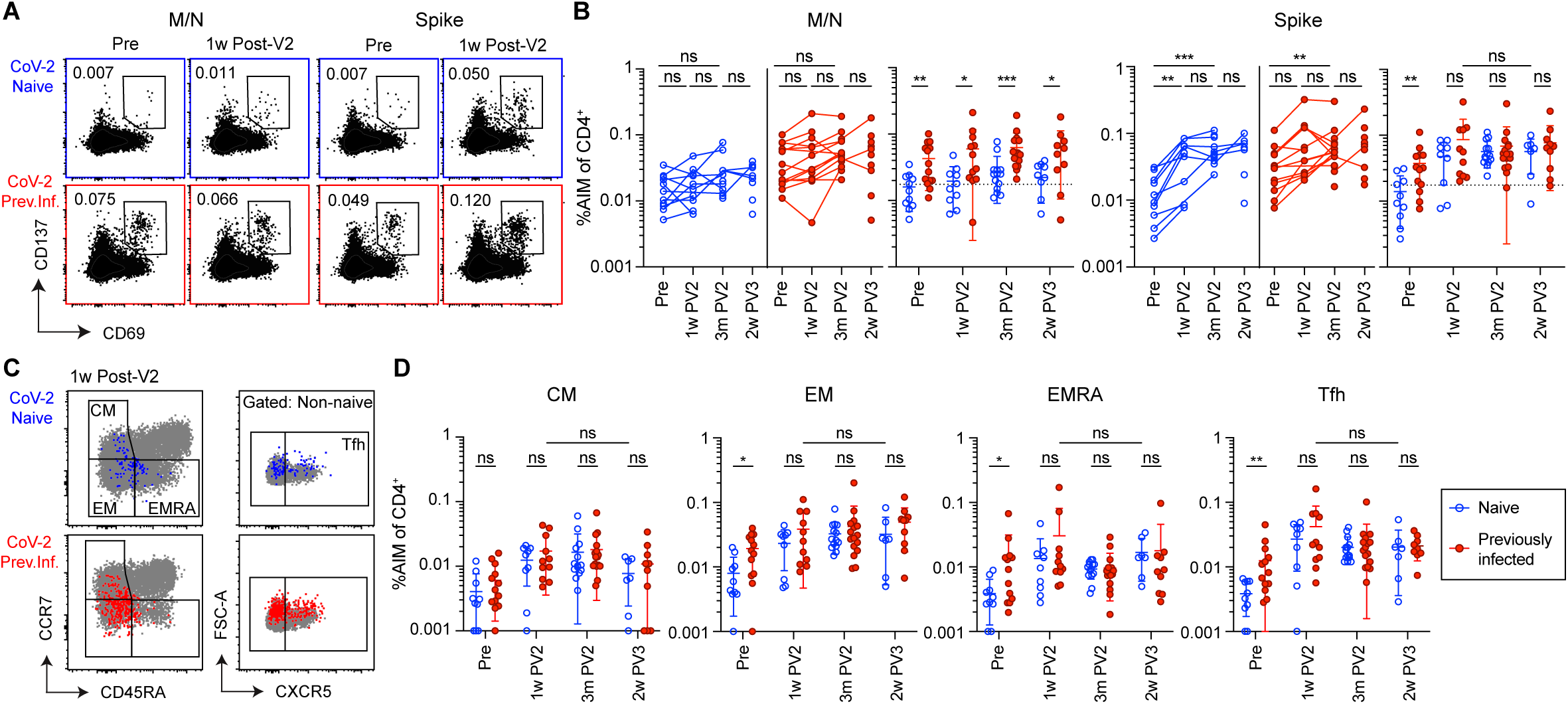
Robust and durable CD4+ T cell responses to SARS-CoV-2 in both previously naive and previously infected individuals. Analysis of non-naive CD4+CD69+CD137+ T cells (AIM CD4+) in SARS-CoV-2 Naive (blue) and Previously Infected (red) individuals. (A and B) Representative flow cytometry plots and summary graphs for total AIM CD4+ T cells. T cell responses shown are either to SARS-CoV-2 Membrane and Nucleocapsid (M/N) or Spike peptide pools for each donor. Data in (B) are represented both longitudinally (left) and by cross-group comparisons (right). (C and D) Representative flow cytometry plots and summary graphs of indicated AIM CD4+ memory and Tfh subsets for donors shown in (A) and (B). Significance was determined by Wilcoxon matched-paired signed rank test for longitudinal analyses and multiple unpaired Mann-Whitney test for group analyses: \**P* < 0.05, \*\**P* < 0.01, \*\*\**P* < 0.001, and \*\*\*\**P* < 0.0001. Error bars represent mean and SD. Dashed lines indicate average donor background level. Lines connecting data points indicate paired samples from the same donor. Central memory (CM), effector memory (EM), TEMRA (T effector memory CD45RA+), Tfh (T follicular helper). Pre-vaccination (Pre), one week post two-dose COVID-19 mRNA vaccination (1w PV2), three months post two-dose vaccination (3m PV2), two weeks post third vaccination dose (2w PV3). See also: Figures S2-S4 and Supplementary Table 2.

We next examined the relative composition of effector and memory populations within the SARS-CoV-2-reactive T cell compartment in N and PI subjects. Prior to vaccination, there was no difference in the frequency of S-reactive central memory (CM) T cells between N and PI groups, however PI individuals maintained significantly elevated frequencies of effector memory (EM and EMRA) and follicular helper (Tfh) cells (**Figures 2C, 2D**). Two-dose vaccination induced a significant increase in nearly all AIM+ effector and memory populations in both groups, with EM and Tfh populations dominating the response (**Figure S4A**). In addition, two-dose vaccination was sufficient to normalize numerical differences identified in pre-vaccination N and PI donors, while a third dose was not associated with any additional increase in these populations in either N or PI donors (**Figure 2D** and **S4A**). Therefore, vaccination induces numerically equivalent S-specific memory Tfh and EM CD4+ T cell responses in N and PI individuals that do not further increase in response to an additional vaccine dose.

### Infection prior to vaccination corresponds with a sustained CD21^-^CD11c^+^ activated B cell response and superior SARS-CoV-2 variant-neutralizing antibody

Memory B cells function by activating, proliferating and forming antibody-secreting plasmablasts more rapidly than naive B cells (Cancro and Tamayko, 2021). Prior to vaccination, RBD-specific B cells in N individuals were predominantly Naive (CD21^+^CD27^-^) while RBD-specific B cells in PI individuals were predominantly non-Naive antigen-experienced B cells (CD21^+^CD27^+^ and CD21^-^CD27^+/-^)(**Figures S5A and S5B**). Two vaccinations induced dominant populations of activated MBCs (CD21^-^CD27^+/-^CD11c^+^)(Cancro and Tomayko, 2021) in both groups at 1 week post-vaccination (**Figures 3A and 3B**). By 3 months post-vaccination, RBD-specific antigen experienced cells in both groups were enriched for IgG^+^ Classical MBCs (cMBCs, CD21^+^CD27^+^) (**Figure 3C**), which can rapidly form IgG-secreting plasmablasts upon antigen exposure (Laidlaw and Ellebedy, 2021) However, a significantly greater population of activated MBCs remained in PI individuals at this memory time point than in N individuals. This may reflect prolonged antigen retention and recent egress of memory B cells from an ongoing germinal center (GC) which would contribute to the increased quantity of RBD-specific antigen-experienced cells including cMBCs and long lived plasma cells producing RBD-specific antibodies in PI individuals with hybrid immunity (**Figures 1C-E**) (Betts et al., 2017; Kim et al., 2019). While mucosal infections such as SARS-CoV-2 are associated with IgA^+^ MBCs particularly at the site of infection, PI individuals had only a small proportion of circulating IgA+ RBD-specific antigen-experienced B cells at 9 months post-infection and they were not further induced in N or PI individuals by vaccination (**Figure S5C**).

**Figure 3.**
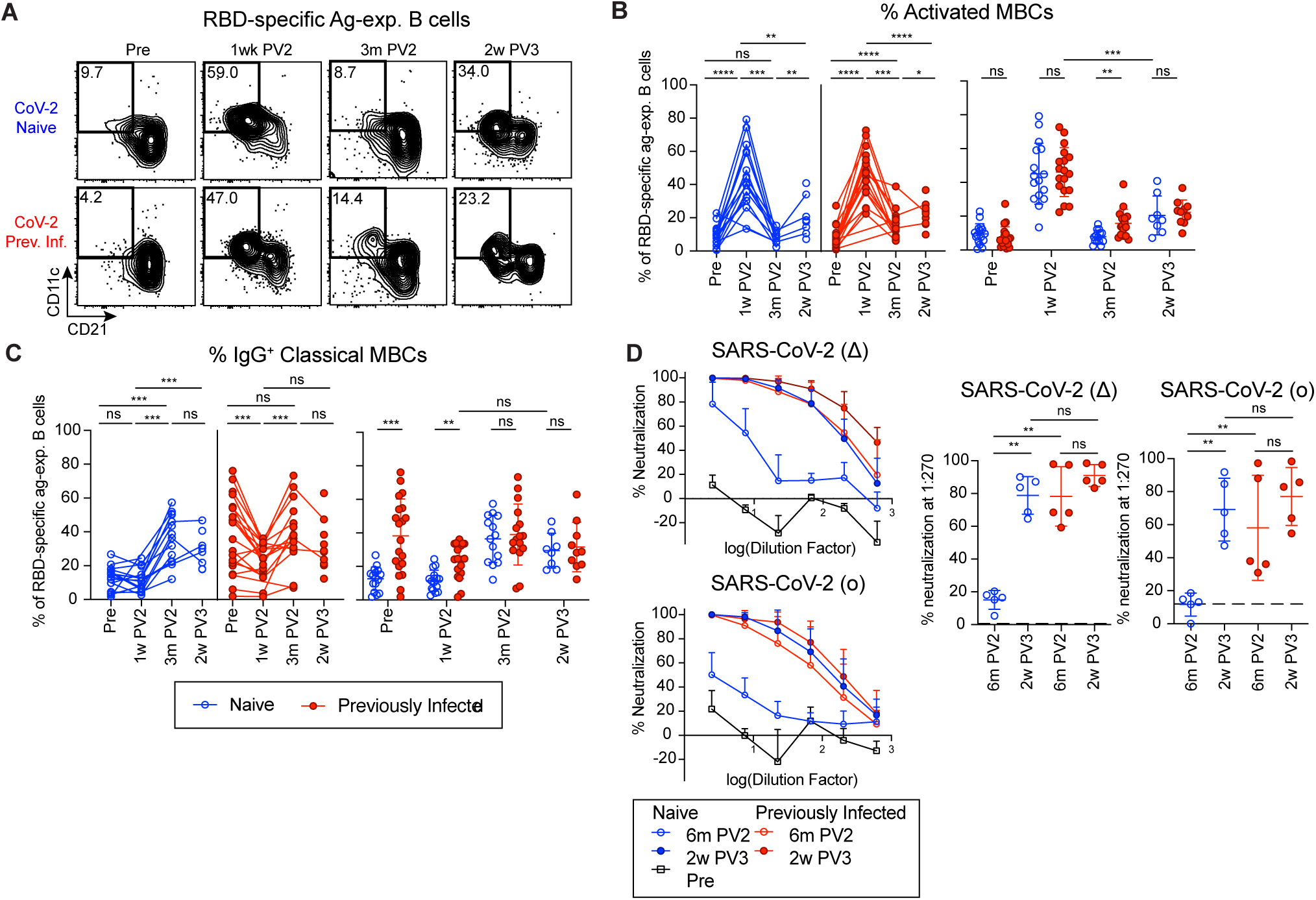
Infection prior to vaccination corresponds with a sustained CD21^-^CD11c^+^ activated B cell response and superior SARS-CoV-2 variant-neutralizing antibody. (A) Representative gating on RBD-specific antigen-experienced (ag-exp.) B cells for activated MBCs (CD21^-^CD11c^+^) from N and PI PBMCs at the indicated time points. (B) Percent of activated MBCs and (C) percent of IgG+ classical MBCs (CD21^+^CD27^+^) of RBD-specific ag-exp. B cells in N and PI PBMCs at the indicated time points. (D) Percent neutralization of SARS-CoV-2(Δ) and SARS-CoV-2(o) spike-pseudotyped virus by plasma from N and PI individuals at the indicated time points (left). Percent neutralization at 1:270 plasma dilution (right). Dashed line indicates the average % neutralization of plasma from N individuals pre-vaccination. Statistics determined by 2-tailed Mann-Whitney tests for all tests because of the limited pairing with 2w PV3: not significant (ns), \**P* < 0.05, \*\**P* < 0.005, \*\*\**P* < 0.0005, and \*\*\*\**P* < 0.0001. Error bars represent mean and SD. Pre-vaccination (Pre), one week post two-dose COVID-19 mRNA vaccination (1w PV2), three months post two-dose vaccination (3m PV2), two weeks post third vaccination dose (2w PV3). See also: Figure S5.

To assess if infection prior to vaccination impacts the breadth or ability of SARS-CoV-2 specific MBCs and antibodies to recognize and combat SARS-CoV-2 variants of concern (VOCs), we measured the ability of RBD-specific MBCs and antibody to recognize the B.1.351 (β) variant and plasma antibody to neutralize the B.1.617.2 (Δ) and B.1.1.529 (o) variants. SARS-CoV-2(β) and SARS-CoV-2(o) have been found to significantly reduce neutralization by vaccine-induced antibodies as compared to the original SARS-CoV-2(Wu-1) strain which was also used to make the mRNA vaccines relevant to this study (Cameroni et al., 2021, Collier et al., 2021). Using SARS-CoV-2 RBD(Wu-1) and SARS-CoV-2 RBD(β) tetramers simultaneously (**Figure S1B**), we found that the majority of RBD-specific antigen-experienced B cells in all N and PI participants could recognize both variants (**Figures S5D** and **S5E**). This technique does not allow us to distinguish cross-reactive cells that bind epitopes shared by the variant RBDs from cells that bind epitopes with the 3 amino acid differences between the variants. While vaccination induced increased RBD(β)-specific IgG antibody titers in both groups, PI individuals had significantly higher RBD(β)-specific IgG titers than N individuals pre- and 6 months post-vaccination (**Figure S5F**). Additionally, we found that plasma from PI individuals 6 months post-vaccination was significantly better at neutralizing SARS-CoV-2(Wu-1) virus as well as pseudotyped SARS-CoV-2(Δ)-spike and SARS-CoV-2(o)-spike lentiviruses than plasma from N individuals (**Figures 3D** and **S5G**). SARS-CoV-2(Wu-1) neutralization was highly correlated with RBD(Wu-1)-specific IgG antibody titers as we and others have shown previously and likely contributes to the improved breadth in PI individuals post-vaccination (**Figure S5H**)(Rodda et al., 2021; Goel et al., 2021). The increased antibody response to a third vaccine dose in N individuals (**Figures 1D, 1E and S5F**) correlated with sharply increased neutralization capacity of the plasma antibody against the variants tested reaching the levels in PI individuals pre- and post-3rd vaccination (**Figures 3D** **and S5F**). Thus, vaccination induces robust humoral memory in N and PI participants, but a third dose is needed for N individuals to achieve the breadth and VOC neutralizing efficacy of the plasma antibody formed in hybrid immunity.

### Infection induces functionally distinct SARS-CoV-2-specific CD4+ T cell responses that are not attributed solely to the number of antigen exposures

During T cell priming, the context of the initial antigen exposure directs CD4+ T cells towards different helper and memory fates with distinct functional capacities. To determine how a primary infection versus vaccination or total number of antigen exposures differentially impacted functional outcomes, we compared the CD4+ T helper lineage composition of SARS-CoV-2 AIM+ T cells based on differential chemokine receptor expression in N and PI subjects. Prior to vaccination, we found that S-reactive T cells in PI subjects were primarily Th1 and Th1/17 memory cells (**Figure 4A** and **4B**). In response to vaccination, the frequency of nearly all T helper subsets examined were increased in both N and PI individuals (**Figure S6A**), consistent with previous reports (Goel et al., 2021; Guerrera et al., 2021; Painter et al., 2021). However, PI subjects maintained an elevated frequency of both CXCR3+CCR6- Th1 and CXCR3+CCR6+ Th1/17 cells, whereas N donors showed increased numbers of CXCR3-CCR6-CCR4+ Th2 cells by 3 months post-vaccine two (**Figure 4B**). Th1 and Th1/17 AIM+ cells continued to trend higher in PI individuals following a third vaccine dose, but this difference did not reach statistical significance. In addition, at this acute timepoint post-third vaccine, the frequency of Th2 populations were normalized between groups (**Figure 4B**). These data demonstrate that T helper polarization in response to initial vaccination differs according to prior SARS-CoV-2 infection status, and that additional vaccine doses may have a normalizing effect on some aspects of this response.

**Figure 4.**
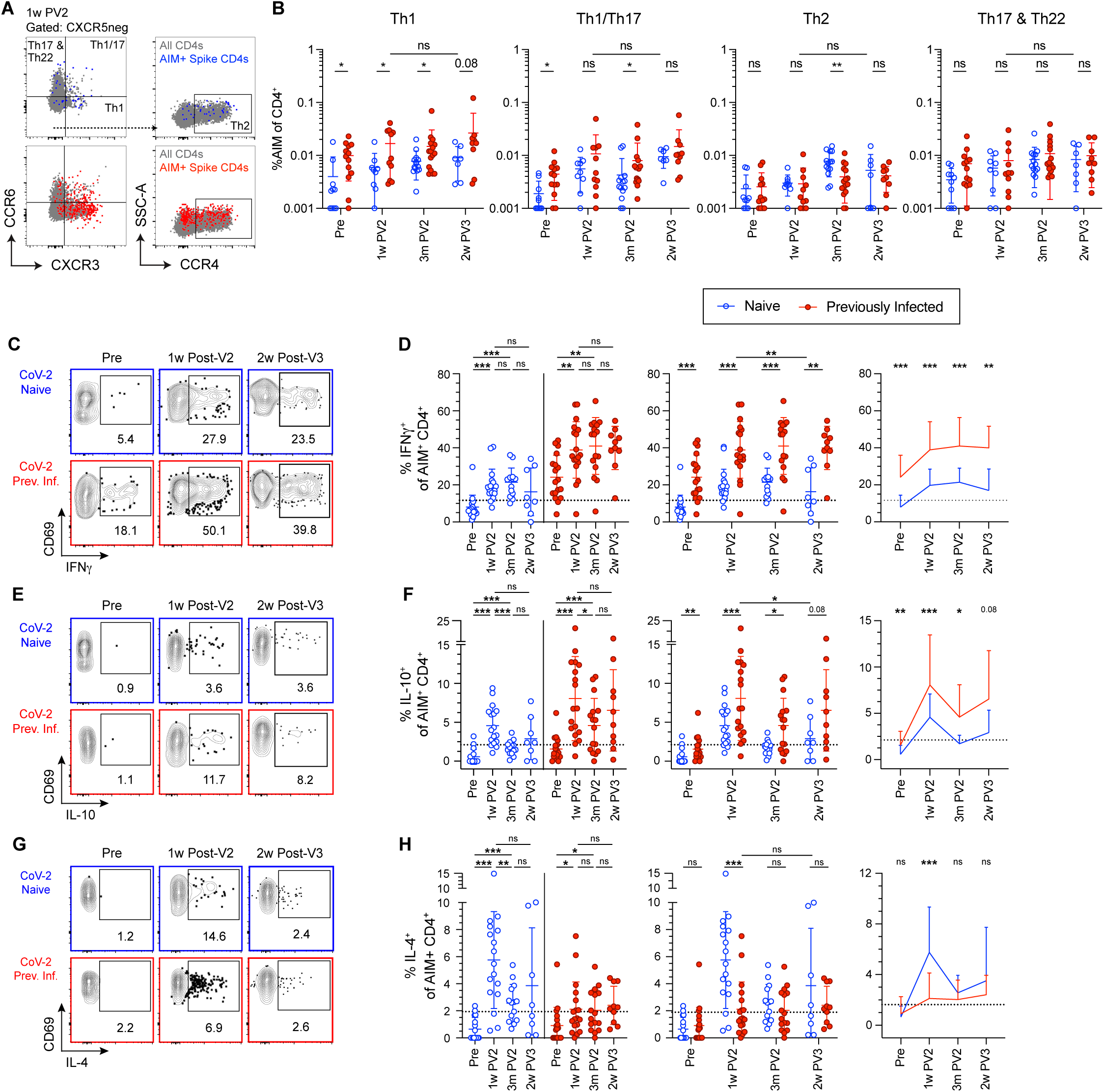
CD4+ T cell responses to SARS-CoV-2 demonstrate qualitative effector cell differences between previously naive and previously infected individuals. Analysis of spike-reactive CD4+ T cells in SARS-CoV-2 Naive (blue) and Previously Infected (red) individuals. (A and B) Representative flow cytometry plots, gating scheme, and summary graphs for non-naive CD4+CD69+CD137+ T cells (AIM CD4+) for the indicated T helper subsets. (C thru H) Representative flow cytometry plots and summary graphs for non-naive CD4+CD69+CD154+ T cells (AIM CD4+) for the indicated cytokines. Data in (D), (F), and (H) are represented both longitudinally (left) and by cross-group comparisons (middle, right). Significance was determined by Wilcoxon matched-paired signed rank test for longitudinal analyses and multiple unpaired Mann-Whitney test for group analyses: \**P* < 0.05, \*\**P* < 0.01, \*\*\**P* < 0.001, and \*\*\*\**P* < 0.0001. Error bars represent mean and SD. Dashed lines indicate average donor background level. Pre-vaccination (Pre), one week post two-dose COVID-19 mRNA vaccination (1w PV2), three months post two-dose vaccination (3m PV2), two weeks post third vaccination dose (2w PV3). See also: Figures S6 thru S11.

To complement the phenotypic analysis of CD4+ T cells, we employed a similar assay to directly interrogate cytokine production from SARS-CoV-2-specific CD4+ T cells following stimulation with M/N or S peptide pools. As demonstrated previously, CD154 and CD69 can be used to identify activated, antigen-reactive T cells (Reiss et al., 2017) that represent the primary cytokine producers in response to stimulation (**Figures S7A** and **S7B**). Importantly, the response measured in CD154+CD69+ SARS-CoV-2 reactive T cells to peptide stimulation mirrors that of AIM+ CD69+CD137+ cells shown in Figure 2, including an M/N response exclusive to PI subjects and robust induction of S-specific cells following two-dose immunization that was maintained after a third vaccination dose in both groups (**Figures S8A-S8C**).

In PI subjects, we measured a strong cytokine response to infection in S- (**Figures 4C-4F, S9A**, and **S9B**) and M/N-reactive T cells (**Figures S10A-E**) that was dominated by the production of IFN-γ and IL-2, but also included modest levels of IL-10. Cytokine production in response to M/N peptides in PI individuals was not enhanced by vaccination (**Figures S10A-E**). Neither N nor PI donor S-reactive T cells produced any detectable IL-4 (**Figures 4G** and **4H**), IL-21, IL-13, IL-17, CD107a, or IL-22 (**Figures S9C**-**S9L**) prior to vaccination. The response in N individuals to two-doses of vaccination generated a functionally diverse CD4+ T cell response, exhibiting strong induction of IFN-γ, IL-10, IL-4 (**Figures 4C-4H**), IL-2, and IL-21 (**Figures S9A-D**), while we observed no increased expression of IL-17A, CD107a, or IL-22 over pre-vaccination levels **(Figures S9F-H)**. IL-4 production, a hallmark of both Th2 and Tfh CD4+ T cell responses (Ruterbusch et al., 2020), was enriched specifically at one week post-vaccination in N individuals (**Figures 4G** and **4H**). However, few spike-reactive T cells co-produced IL-4 and IL-13 **(Figure S9E)**, suggesting the absence of bonafide Th2 polarization. Consistent with our phenotypic analysis of T helper polarization based on differential chemokine receptor expression, PI individuals not only exhibited a strong induction of IFN-γ-producing S-reactive CD4+ T cells following vaccination, but this response remained significantly elevated compared with the N group following both second and third vaccine doses (**Figures 4C** and **4D**). PI donor CD4+ T cells also produced higher levels of IL-10 after two-dose vaccination compared with N donors, which also trended higher after a third vaccine dose despite failing to reach statistical significance (**Figures 4E** and **4F**). Production of IL-21, although not detected following infection, was observed at comparable levels in both N and PI individuals following two- and three-dose vaccination (**Figures S9C** and **S9D**).

The S-reactive CD4+ T cell compartment was further distilled into subpopulations using a dimensionality reduction analysis on pooled CD154+CD69+ cells from all donors. UMAP followed by FlowSOM analysis identified 15 distinct clusters, of which 13 were expanded following infection or vaccination, and 6 included cells which produced cytokine(s) (**Figures S11A-P**). Two of these clusters, CL4 and CL13, were specifically enriched in PI donor T cells following vaccination. CL4 was defined largely by the production of IFN-γ and low CD127 expression, indicative of terminally differentiated Th1 cells (**Figure S11E**). CL13 was defined by co-production of IFN-γ and IL-10, while also displaying low CD127 expression, resembling FOXP3-type-1 regulatory (Tr1) cells (**Figure S11N**) (Jankovic et al., 2007; Sun et al., 2009) In contrast, increased vaccine-induced T cells producing IL-4 were specifically enriched in CL15 only in N donors. These cells additionally co-produced IL-2, IFN-γ, and small amounts of IL-21 (**Figure S11P**), suggesting a Tfh-like phenotype (Olatunde et al., 2021). Collectively, this unbiased analysis of the S-reactive CD4 T cell compartment identifies the emergence of functionally distinct subsets which differ following vaccination in PI and N participants according to prior SARS-CoV-2 infection history.

To determine how the number of antigen exposures may alter distinct cytokine production profiles seen in N and PI individuals, we compared S-reactive T cells from N individuals after three doses of vaccination (3 antigen exposures) to those from PI individuals after two doses of vaccination (3 antigen exposures). A comparison of AIM+ S-specific CD4+ T cells in triple vaccinated N subjects to double vaccinated PI individuals normalized differences between Th1 and Th1/17 frequencies as assessed by CXCR3 and CCR6 expression (**Figures 4A** and **4B**). We also found similar frequencies of CM, EM, EMRA and and Tfh cells when normalizing for the number of antigen exposures (**Figures 2B** and **2D**). However, when cytokine production was assessed between triple vaccinated N donors and double vaccinated PI donors, the PI group maintained a dramatically elevated frequency of IFN-γ- and IL-10-producing S-specific CD4+ T cells compared with the N group (**Figures 4C** and **4D**). These data indicate that T cell priming by infection or vaccination promotes distinct effector states in which phenotypic outcomes can be normalized by additional antigen exposure, while functional differences persist through multiple vaccine doses.

## Discussion

Protective immunity generated in response to infection or vaccination is characterized by an expanded population of antigen-specific memory cells that rapidly expresses effector molecules upon antigen re-exposure. In order to prevent or control reinfection, memory cells are functionally tailored by the initial priming event, however it is unclear how subsequent antigen exposures alter the quantity and quality of the response. The emergence of a novel coronavirus and expeditious generation of vaccines provided an unprecedented opportunity to examine the formation of a nascent antigen-specific memory pool in human volunteers, and the impact of previous infection on the quantity, quality and durability of vaccine-induced immune memory. This is particularly important as SARS-CoV-2 infection prior to vaccination, referred to as “hybrid immunity”, provides a protective advantage over vaccination alone (Abu-Raddad et al., 2021; Crotty 2021; Gazit et al., 2021; Goldberg et al., 2021).

In this study, we examined the distinguishing features of hybrid immunity to SARS-CoV-2 in comparison to vaccination alone over time and with subsequent antigen exposures. We found that certain features of the humoral response differed in N and PI subjects following vaccination that could contribute to hybrid immunity. These include the enhanced numbers of RBD-specific memory B cells and SARS-CoV-2 variant-neutralizing plasma antibody. These features were normalized following booster vaccination, and therefore could be a function of the number of antigen exposures. However the cytokine profiles of spike-specific memory CD4+ T cells in previously infected and then vaccinated individuals exhibited a unique IFN-γ and IL-10 cytokine-producing profile that could not be achieved by multiple vaccinations in the absence of prior infection, and this represents the first immune correlate of hybrid immunity that is stably imprinted on responding cells during initial infection.

Enhanced numbers of memory B cells and breadth of antibody specificity are proposed to be essential to the mechanisms underlying the superior immunity described in PI individuals after vaccination (PV2) (Cho et al., 2021; Schmidt et al., 2021; Schmidt et al., 2021; Stamatatos et al., 2021; Wang et al., 2021). In accordance with these studies, we found infection prior to vaccination induced higher numbers of RBD-specific MBC, IgG and IgA circulating antibodies, and increased plasma neutralization of multiple VOCs including SARS-CoV-2(o). Furthermore, differences in the number and timing of SARS-CoV-2 antigen exposures impacts memory B cell response maturation (Goel et al., 2021). This is likely to explain the quantitative differences in vaccine-induced humoral memory and antibody breadth between N and PI individuals described in our study because additional vaccination normalized the differences between groups. We additionally observed however that RBD-specific MBCs in PI individuals were enriched for CD21^-^CD11c^+^ activated MBCs 3 months post-vaccination and hypothesize these may be an indicator of ongoing GC responses possibly supported by the enrichment of IFN-γ-expressing CD4+ T cells which can drive T-bet as well as support activation of B cells. CD21-CD11c+ memory cells have been associated with Tbet expression, can serve as potent antigen presenting cells, and have been described in the context of chronic viral infection and autoimmunity (Knox et al., 2017; Glass et al., 2020). Sustained low-levels of activation may enrich the ongoing germinal center responses that have been shown to last for at least 6 months post-vaccination and support generation of higher affinity and broadly neutralizing antibodies critical for combating variants (Cho et al., 2021; Kim et al., 2021; Turner et al., 2021) and likely contributing to the protective advantage of hybrid immunity. Additional data from memory time points after third vaccination are needed to determine whether this enhanced activation at memory time points can be matched by repeat vaccination.

SARS-CoV-2 infection and COVID-19 immunization both induce robust Th1 immune memory as found here and by other groups (Goel et al., 2021; Guerrera et al., 2021; Painter et al., 2021), but the CD4+ T cell cytokine profiles induced by infection are distinct and not recapitulated by repeat vaccination. Mucosal infection with SARS-CoV-2 virus and intramuscular immunization with full-length spike mRNA induce distinct environments for immune activation to spike protein. Viruses in the *coronaviridae* family antagonize host production of type-I and -III interferon in response to pattern recognition receptors (PRRs) (Lei et al., 2020; Liu et al., 2021; Liu et al., 2021; Thoms et al., 2020; Xia et al., 2020). The inability of infected cells to sense viral PAMPs and upregulate antiviral programs permits prolonged viral replication in pulmonary tissue and has been associated with sustained and dysregulated production of the inflammatory cytokines IL-6, IL-8, TNF-α, and IL-12 in COVID-19 patients (Galani et al., 2021; Lucas et al., 2020). In contrast, mRNA vaccination provides a coordinated and succinct innate immune response, with reduced levels of inflammatory cytokines during the period of T cell differentiation and/or contraction into memory populations. Prolonged inflammatory cytokine production by innate immune cells following SARS-CoV-2 infection may induce the CD4+ memory T cell IFN-γ and IL-10 signature we describe in PI individuals that is maintained through vaccination. The inflammatory cytokines IL-12, IFN-α, and IFN-γ, produced by innate lymphocytes in response to viral PAMPs, directly drive T cells to upregulate the transcription factor T-bet, the master transcription factor for Th1 cells (Brinkmann et al., 1993; Lazarevic et al., 2013). Further, the cytokines IL-6, IL-12, or IL-27 drive the expression of IL-10 in FOXP3-CD4+ helper T cells through the adapter c-maf (Gabryšová et al., 2018; Pot et al., 2009; Yang et al., 2005). Finally, infection with SARS-CoV-2 recruits effector T cells to the lung tissue, generating bona fide tissue-resident memory (Trm) cells in the human lung (Poon et al., 2021). Therefore, S-reactive CD4+ T cells from PI individuals may receive additional tissue-restricted signals, before contracting into memory cells (Klicznik et al., 2019; Wijeyesinghe et al., 2021). Indeed, Trm cells produce the highest levels of IFN-γ and IL-10 in a mouse-adapted SARS-CoV infection (Zhao et al., 2016). Therefore, it may also be the case that the increase in cytokine production we see in PI donors is due to the presence of circulating S-reactive T cells which previously received tissue-restricted cues.

The distinct CD4+ T cell cytokine signature we describe likely contributes to the increased protection observed in PI individuals with hybrid-immunity. IFN-γ is produced rapidly by memory T cells *in vivo*, driving an antiviral response program which is characterized by the upregulation of interferon-stimulated genes. Our data showing elevated frequencies of IFN-γ-producing memory CD4+ T cells in PI individuals and the observed localization of CD4 Trm in the lung (Poon et al., 2021) together support a model in which early antiviral programs are bolstered rapidly by the inclusion of memory T cells locally at the site of infection. The ability of memory T cells to detect presented SARS-CoV-2 antigens may be especially important in the context of CoVs, which repress canonical innate immune PRRs. Indeed, pre-treatment of naive mice with rIFN-γ completely inhibits SARS-CoV infection of the lung (Zhao et al., 2016). This study also reported that concomitant treatment with rTNFα exacerbates SARS-CoV-mediated disease in mice. IL-10 can inhibit TNFα and other pro-inflammatory cytokines in order to regulate inflammatory responses to infection (De Waal Malefyt et al., 1991; Gazzinelli et al., 1996; Hunter et al., 1997). Therefore, increased production of IL-10 by S-specific Th1 cells potentiated through viral infection could be a critical element of hybrid immunity that limits future symptomatic SARS-CoV-2 infection. Additional work is needed to explore this crucial role of memory T cell-derived IL-10 in secondary challenge.

The protective advantage of hybrid immunity likely stems from a combination of higher numbers of SARS-CoV-2-specific memory B cells, higher neutralizing antibody titers and the infection-imprinted IFN-γ and IL-10 cytokine profile in CD4+ T cells. These features were not as highly induced in solely vaccinated individuals even with a third antigen exposure via vaccination. Future work is needed to determine how the relative timing of infection vs. vaccination quantitatively and qualitatively impacts functional SARS-CoV-2 immune memory. The reduced immune response to the third vaccination in both groups as compared to the first two suggests that further homologous vaccinations at least at these doses are unlikely to bolster SARS-CoV-2 cellular immune memory. Therefore, as additional viral variants evolve to further evade immune memory, we will need to expand the pool of SARS-CoV-2-specific memory cells and increase the titers of viral variant neutralizing antibodies. To do this, additional vaccine doses likely need to include variant spike proteins to engage a broader repertoire of immune cells and induce further cross-reactive memory responses.

## Data Availability

All data generated or analysed during this study are included in this published article (and its supplementary information files) or available from the corresponding author upon reasonable request with the exception of a few blood draw samples that were used up in this study.
Correspondence and requests for materials should be addressed to M.P.

## Experimental Methods

### Ethics Statement

Participants were enrolled in the Hospitalized or Ambulatory Adults with Respiratory Viral Infections (HAARVI) study (STUDY00000959), Healthy Adult Specimen Repository study (STUDY00002929) or COVID-19/SARS-CoV-2 Prevalence and Antibody Therapy Development study (Gale Lab, STUDY00009810). All studies are approved by the University of Washington Human Subjects Division Institutional Review Board. Informed consent was obtained from all enrolled participants. Samples were de-identified prior to analysis.

### Study participants and clinical data collection

Participants were enrolled in one of three prospective cohort studies for longitudinal tracking of immune responses to COVID-19 vaccination in WA, USA. Previously SARS-CoV-2 infected (PI) individuals reported a positive SARS-CoV-2 PCR nasal swab or in one case a positive antibody test within 3 months of symptom onset in 2/2020-10/2020. PI individuals reported mildly symptomatic disease not requiring hospitalization. Participants were considered SARS-CoV-2 naive (N) prior to vaccination based on no prior positive SARS-CoV-2 PCR nasal swab and no detectable SARS-CoV-2 RBD IgG by ELISA (below a threshold of mean + 3 standard deviations of historical negative plasma samples drawn prior to 2020). All participants completed surveys regarding symptom and demographic information. Blood draws were collected pre-vaccination (N: 21, PI: 23), 1 week post-vaccination two (N: 18, PI: 22), 3 months post-vaccination two (N: 15, PI: 23), 6 months post-vaccination two (N: 8, PI: 9) and 2 weeks post-vaccination three (N: 8, PI: 10). Not all samples were run for both plasma and PBMC assessments.

### Sample processing and plasma collection

Venous blood from study volunteers was collected in EDTA tubes and spun at 700xg for 10 min. Plasma was collected, heat-inactivated at 56°C for 30 min and stored at -80°C. Cellular fraction was resuspended in phosphate buffered saline (PBS) and PBMCs were separated from red blood cells using Sepmate PBMC Isolation Tubes (STEMCELL Technologies) according to manufacturer’s instruction or cells were resuspended in HBSS, washed, overlaid with ficoll, spun at 400xg for 30 min with no brake and PBMCs collected. Cells were washed twice in HBSS and frozen at -80°C before being stored in liquid nitrogen.

### SARS-CoV-2 RBD protein and tetramer generation

Recombinant SARS-CoV-2 RBD (Wuhan-1, Wu-1) was generated by standard transient transfection followed by IMAC purification as described previously (Walls et al., 2020). Recombinant SARS-CoV-2 RBD (B.1.1.351, β) was generated by transient transfection from the SARS-CoV-2-Beta-RBD-Avi (K417N-E484K-N501Y) construct synthesized by GenScript into CMVR with an N-terminal mu-phosphatase signal peptide and a C-terminal octa-histidine tag, flexible linker, and avi tag (GHHHHHHHHGGSSGLNDIFEAQKIEWHE) as described previously (Tortorici et al., 2021). For tetramer generation, RBD proteins were biotinylated with the BirA500 kit (Avidity), tetramerized with streptavidin-phycoerythrin (SA-PE) or streptavidin-allophycocyanin (SA-APC) (Agilent) and stored in 50% glycerol at -20°C as previously described (Krishnamurty et al., 2016). Decoy reagents were generated by tetramerizing an irrelevant biotinylated protein with SA-PE previously conjugated to Dylight594 NHS Ester (ThermoFisher) and Dylight650 NHS Ester (ThermoFisher) or SA-APC previously conjugated to Dylight755 NHS Ester (ThermoFisher).

### ELISA

96-well plates (Corning) were coated with 2 ug/mL of recombinant SARS-CoV-2 RBD or trimeric S protein diluted in PBS and incubated at 4°C overnight. Plates were washed with PBS-T (PBS containing 0.05% Tween-20) and incubated with blocking buffer (PBS-T and 3% milk) for 1h at room temperature (RT). Plasma, culture supernatants or monoclonal antibodies were serially diluted in dilution buffer (PBS-T and 1% milk) in triplicate, added to plates, and incubated at RT for 2 h. Secondary antibodies were diluted in dilution buffer as follows: anti-human IgG-HRP (Jackson ImmunoResearch) at 1:3000, anti-human IgM-HRP (Southern Biotech) at 1:3000, or anti-human IgA-HRP (Southern Biotech) at 1:1500. Plates were incubated with secondary antibodies for 1h at RT, then detected with 1X 3,30,5,50-Tetramethylbenzidine (TMB) (Invitrogen) and quenched with 1M HCl. Sample optical density (OD) was measured by a spectrophotometer at 450nm and 570nm.

### Generating pseudotyped lentiviral particles with SARS-CoV-2 spike protein

The SARS-CoV-2(o)-spike pseudotyping plasmid was assembled by introducing gene fragments synthesized by Integrated DNA Technologies (Coralville, IA, USA) encoding the SARS-CoV-2(o) variant (B.1.1.529, omicron) spike gene to replace the Wu-Spike_D614G sequence in a digested pseudotyping plasmid backbone (BEI Resources; NR-53765) using standard molecular biology techniques, including NEBuilder HiFi DNA Assembly (New England Biolabs, Ipswich, MA, USA). Plasmid sequence was confirmed by Sanger sequencing. The SARS-CoV-2(o)- and SARS-CoV-2(Δ)-spike pseudotyped lentiviral vectors (LVs) were each produced by transient polyethylenimine transfection of 293T cells with a CMV.Luc.IRES.GFP vector plasmid (BEI Resources; NR52516), a second generation helper plasmid pMD2.g (gift from Didier Trono; Addgene #12260), and either: the SARS-CoV-2(o) pseudotyping plasmid above, or a plasmid encoding the SARS-CoV-2(Δ) variant spike (Invivogen, San Diego, CA,USA; pLV-Spike-V8, B.1.617.2, delta). At 24 hours following transfection, cell culture media was exchanged for fresh DMEM with 3% FCS. Supernatants were harvested the next day, filtered through 0.2uM filters, and concentrated by overnight centrifugation at 4°C. Pellets were then resuspended in PBS at 100X.

### Pseudovirus neutralization test (pVNT)

PVNT assays were performed as previously described (Crawford et al., 2020). Briefly, heat inactivated plasma was diluted 1:5 followed by four 3-fold serial dilutions all in duplicate and mixed 1:1 with 10^6^ relative luciferase units of SARS-CoV-2(Δ)-spike or SARS-CoV-2(o)-spike pseudotyped lentivirus in DMEM with 10% heat-inactivated FBS, 2 mM L-glutamine, 100 U/mL penicillin, and 100 µg/mL streptomycin. After 1 hour incubation at 37°C, the plasma/virus mixtures were added to 96-well poly-L-lysine-coated plates seeded with human ACE2-expressing 293T cells (BEI Resources: NR-52511) 20 hours prior. Each plate contained wells with no plasma and wells with no plasma and 293T cells as a background control. Human ACE2-Fc (BEI Resources) was used as a positive control with a starting dilution at 10ug/ml. After incubating for 48 hours, supernatant was pipet off and replaced with Bright-Glo Luciferase Assay System luciferase (E2610, Promega) for 2 min at 25°C in the dark before transferring to black-bottom plates for measuring luminescence for 1s per well on a Centro LB 960 Microplate Luminometer (Berthold Technologies). Percent neutralization was calculated as (1 – (sample+293T-ACE2+virus RLU - 293T+virus RLU)/(293T-ACE2+virus RLU - 293T+virus RLU) x 100. Data was collected with MikroWin2000 and analyzed in Prism (GraphPad).

### Plaque reduction neutralization test (PRNT)

PRNT assays were performed as previously described (Erasmus et al., 2020). Briefly, heat inactivated plasma was diluted 1:5 followed by four 4-fold serial dilutions and mixed 1:1 with 600 PFU/mL SARS-CoV-2 WA-1 (BEI resources) in PBS+0.3% cold water fish skin gelatin (Sigma). After 30 min of incubation at 37°C, the plasma/virus mixtures were added to 12 well plates of Vero cells and incubated for 1 h at 37°C, rocking every 15 min. All dilutions were done in duplicate, along with virus only and no virus controls. Plates were then washed with PBS and overlaid with a 1:1 mixture of 2.4% Avicel RC-591 (FMC) and 2X MEM (ThermoFisher) supplemented with 4% heat-inactivated FBS and Penicillin/Strep-tomycin (Fisher Scientific.) After a 48h incubation, the overlay was removed, plates were washed with PBS, fixed with 10% formaldehyde (Sigma-Aldrich) in PBS for 30 min at room temp and stained with 1% crystal violet (Sigma-Aldrich) in 20% EtOH. Percent neutralization was calculated as (1 – # sample plaques/# positive control plaques) x 100. Data was analyzed in Prism (GraphPad).

### Peptide mesopools

SARS-CoV-2 15-mer peptides, 1mg each (BEI Resources), were provided lyophilized and stored at -80C. Peptides were selected for reactivity against a broad range of class I and class II HLA sub-types for targeted coverage of CD4+ and CD8 T cell epitopes identified previously (Tarke et al., 2021) (**Table S2**). Before use, peptides were warmed to room temperature for 1 hour then reconstituted in DMSO to a concentration of 10 mg/mL. Individual peptides were combined in equal ratios to create Membrane/Nucleocapsid (182ug/mL each, 55 peptides) or Spike (200ug/mL each, 49 peptides) megapools, maintaining a total concentration of 10mg/mL.

### Activation induced marker assay

Approximately 20e6 PBMC from SARS-CoV-2 naive or previously infected individuals were divided in two for full phenotypic or cytokine analysis. For broad surface phenotyping 10e6 PBMC per sample were further divided into four 5mL polystyrene tubes and cells were pelleted at 250xg for 5 minutes. Pellets were resuspended at 5e6/mL in one of the following treatment conditions: DMSO (Sigma-Aldrich, >99.5% cell culture grade), 1ug/mL CEFX Ultra SuperStim Pool (JPT, PM-CEFX-2), or 5ug/mL SARS-CoV-2 Membrane and Nucleocapsid or Spike peptide megapools. Stimulation was performed for 18 hours in ImmunoCult-XF T cell Expansion Medium (StemCell Technologies). For intracellular cytokine assessment 2e6/mL PBMC were stimulated using 6.6ug/mL SARS-CoV-2 peptide megapools, or an equivalent volume of DMSO (Sigma-Aldrich, >99.5% cell culture grade) for 12 hours in RPMI complete T cell medium containing anti-human CD40 antagonist mAb (Miltenyi, clone HB10), 5uL LAMP-1 BV510 mAb (BioLegend, clone H4A3), 2uM CaCl2, and 1.8uL Monensin (Becton Dickinson) - for the final 8 hours of culture.

### Immunophenotyping of PBMCs

PBMCs were thawed at 37°C and washed twice before first staining with decoy tetramer and then with RBD tetramer prior to incubation with anti-PE magnetic beads and magnetic bead enrichment (Miltenyi Biotec) as previously described (Krishnamurty et al., 2016). Cells in the positive fraction were stained with surface antibodies for B cell phenotypes (**Table S3**). Subsequent analysis of T cells was performed on unlabeled PBMC flow-thru resulting from B cell RBD-tetramer pulldown.

PBMC flow-thru were stimulated with peptide megapools as described above. For surface phenotyping, cells were washed and barcoded using four different fluorescently labeled CD45 antibodies to create eight unique barcodes as previously described (Becht et al., 2021). Using this method to limit technical variability, all four stimulation conditions for both pre- and post-vaccination samples were combined and fully stained simultaneously for 20 minutes at 37°C. Cells were then fixed for 10 minutes at room temperature using 1% paraformaldehyde (Sigma-Aldrich). For intracellular cytokine staining, cells were first incubated with anti-CXCR5 antibody at room temperature for 40 minutes. Cells were then stained with surface antibodies for 25 minutes at 4°C and then fixed and permeabilized with Fixation Permeabilization kit (Becton Dickinson) for 15 minutes at room temperature before staining for intracellular cytokines for 20 minutes at room temperature (**Table S3**).

### Fluorescence cytometry

Data were acquired on a five-laser Cytek Aurora (T cell surface phenotyping and T cell intracellular cytokine analysis) or BD FACS Symphony A3 or A5 (B cell surface phenotyping). Control PBMCs or UltraComp eBeads (ThermoFisher) were used for compensation. Up to 10^7^ live PBMC were acquired per sample for T cells and all enriched PBMCs were acquired for B cells. Data were analyzed using SpectroFlow (Cytek Biosciences) and FlowJo10 (Becton Dickinson) software.

### High-dimensional analysis of cytometry data

AIM-positive (CD154+CD69+) cells from all data files were concatenated with keywords and subjected to Phenograph clustering algorithm using k=40 nearest neighbors (Levine et al., 2015) and UMAP dimensionality reduction plugins using parameters IL-2, IFN-γ, IL-10, IL-4, IL-21, CD127, CD25, and CXCR5 in FlowJo 10 (Becton Dickinson). Clusters were then enumerated per 1M non-naive T cells for each sample by multiplying the percentage “cluster X parameter” by percentage “AIM+” and then by 1 million.

### Statistics

Statistics used are described in figure legends and were determined using Prism (Graphpad). All measurements within a group in a panel are from distinct samples. Data is pooled from nine experiments for Pre and 1w PV2, three experiments for 3m PV2, and two experiments for 2w PV3 time points. Statistical significance of all pairwise comparisons was assessed by two-tailed nonparametric tests; Mann-Whitney for unpaired data and Wilcoxon signed rank tests for paired data unless otherwise noted. No multiple hypothesis testing was applied as the metrics tested were selected based on specific hypotheses.

## Acknowledgements

We thank the participants of the HAARVI Research study, Healthy Adult Specimen Repository study and COVID-19/SARS-CoV-2 Prevalence and Antibody Therapy Development study; Megan Kemp and Jen Rathe for sample procurement; Kristin Huden and Callista Nackviseth for sample processing; Jason Netland and Laila Shehata for sample processing and technical help; Lauren Carter (Neil P. King lab) and John Bowen (David Veesler lab) for RBD(Wu-1) and RBD(β) protein; Adam Wojno and the BRI Cell and Tissue Imaging Core and U. Washington Cell Analysis Facility for technical help; Wesley C. Van Voorhis for historical negative plasma samples; Hannah Deberg, Charlie Quinn and Naresh Doni Jayavelu for assistance with statistical analyses; figure cartoon created with BioRender.com; and the Pepper, Campbell and Chu labs for helpful discussion. This work was supported by the following funding: L.B.R. and K.B.P (NIH2T32 AI106677); U. Washington Cell Analysis Facility, Symphony A3 NIH 1S10OD024979-01A1; D.J.C. and P.A.M. (NIH R01AI127726, NIH U19AI125378-S1) and M.P. (NIH U01AI142001-02S1; R01AI118803); BWF #1018486 and COVID Pilot grant to M.P.; and Emergent Ventures Fast Grant to M.P

## Author Contributions

M.P., L.B.R., K.B.P, D.J.C, and P.A.M. conceived the study. M.P., H.C., M.G. and L.B.R. initiated and ran the clinical studies. N.F., J.L. and C.A.H. enrolled participants, managed surveys and coordinated visits. C.S. and I.G. procured samples. C.A.H., L.B.R, N.F. and J.L. processed and preserved blood and plasma samples. L.B.R. generated and validated tetramer reagents. M.H. generated pseudoviruses. L.B.R. and C.A.H. performed ELISA experiments. J.E. performed PRNT experiments. L.B.R. performed and analyzed antigen-specific B cell flow cytometry and pVNT experiments. D.J.C. defined SARS-CoV-2 peptide pool construction. P.A.M. and K.B.P conceived, performed, and analyzed AIM T cell experiments with help from M.F. L.B.R., P.A.M, K.B.P, D.J.C, and M.P drafted the manuscript. All authors helped edit the manuscript. M.P. secured funds and supervised the project.

## Competing Interests

M.P. is a member of the Scientific Advisory Board of VaxArt and NeoLeukin Inc.

## Data and Materials Availability

All data generated or analysed during this study are included in this published article (and its supplementary information files) or available from the corresponding author upon reasonable request with the exception of a few blood draw samples that were used up in this study.

## Supplemental Figure Legends

**Figure S1. COVID-19 vaccination induces SARS-CoV-2 RBD-specific B cell responses in previously infected and naive individuals. Related to Figure 1.**

(A) ELISA area under the curve (AUC) for RBD-specific IgG in plasma collected from individuals prior to 2020 and the SARS-CoV-2 pandemic (historical negatives, HN, black), SARS-CoV-2 naive (N) individuals and previously infected (PI) individuals that tested PCR+ for SARS-CoV-2. Dashed line indicates mean + 3 SD of HN AUC values.

(B) Representative flow cytometry gates for phenotyping RBD(Wu-1)- and RBD(β)- specific B cells from N and PI PBMCs. (C) Representative gating on CD19^+^CD38^lo^RBD-tetramer^+^Decoy^-^ cells for SARS-CoV-2 RBD-specific antigen-experienced (ag-exp.) B cells (CD21^+^CD27^+^ and CD21^-^CD27^+/-^) from N and PI (Prev. Inf.) PBMCs at the indicated time points. (D) Number (left) and percent IgG+ or IgA+ (right) of RBD-specific plasmablasts (PBs, CD27^+^CD38^hi^). Statistics determined by two-tailed Mann-Whitney tests: not significant (ns), \**P* < 0.05, \*\**P* < 0.005, \*\*\**P* < 0.0005, and \*\*\*\**P* < 0.0001. Pre-vaccination (Pre), one week post two-dose COVID-19 mRNA vaccination (1w PV2), three months post two-dose vaccination (3m PV2), two weeks post third vaccination dose (2w PV3). Error bars represent mean and SD.

**Figure S2. Determination of antigen-reactive activation markers in response to SARS-CoV-2 peptide stimulation. Related to Figure 2.**

(A) Representative flow cytometry plots showing gating schematic for non-naive (nn) CD4+ T cells (AIM CD4+). (B) Boolean assessment of activation-associated markers on spike-reactive AIM CD4+ T cells from Naive (blue) and Previously Infected (red) donors comparing pre-vaccine (Pre) and one week post two-dose mRNA vaccination (1w Post-V2). “+”: marker was gated, “-”: marker was excluded, “+/-”: analysis was agnostic to indicated marker. (C) Representative flow cytometry plots for AIM CD4+ CD69+CD137+ T cells. Dot plot overlays show cells expressing additional activation-associated markers: CD134 (Top) and PD-L1 (Bottom). (D) Representative flow cytometry plots for additional AIM markers CD134 and PD-L1. Significance was determined by Wilcoxon matched-paired signed rank test: \**P* < 0.05, \*\**P* < 0.01, \*\*\**P* < 0.001, and \*\*\*\**P* < 0.0001.

**Figure S3. Individual donor background, positive control and numerical quantification of AIM CD4+ T cells. Related to Figure 2.**

(A and B) Representative flow cytometry plots and summary graphs for non-naive CD69+CD137+ T cells (AIM CD4+) from SARS-CoV-2 Naive (blue) and Previously Infected (red) individuals. Data in (B) are to a common antigen control peptide pool (CEFX) and represented both longitudinally (left) and by cross-group comparisons (right). (**C**) Total AIM CD4+ cell count per million CD4+ T cells in each SARS-CoV-2 Naive (left) and Previously Infected (right) donor. (**D**) Background normalization of AIM CD4+ T cell data from (C) was performed by subtracting DMSO cell counts from matched CEFX, M/N, and S samples. Significance was determined by Wilcoxon matched-paired signed rank test for longitudinal analyses and multiple unpaired Mann-Whitney test for group analyses: \**P* < 0.05, \*\**P* < 0.01, \*\*\**P* < 0.001, and \*\*\*\**P* < 0.0001. Error bars represent mean and SD. Dashed lines indicate average donor background level. Pre-vaccination (Pre), one week post two-dose COVID-19 mRNA vaccination (1w PV2), three months post two-dose vaccination (3m PV2), two weeks post third vaccination dose (2w PV3).

**Figure S4. Assessment of AIM CD4+ T cell memory and T follicular helper subsets. Related to Figure 2.**

(**A**) Longitudinal summary graphs of indicated AIM CD4+ memory and Tfh subsets for SARS-CoV-2 Naive (blue) and Previously Infected (red) donors. Significance was determined by Wilcoxon matched-paired signed rank test for longitudinal analyses and multiple unpaired Mann-Whitney test for group analyses: \**P* < 0.05, \*\**P* < 0.01, \*\*\**P* < 0.001, and \*\*\*\**P* < 0.0001. Error bars represent mean and SD. Dashed lines indicate average donor background level. Lines connecting data points indicate paired samples from the same donor. Central memory (CM), effector memory (EM), TEMRA (T effector memory CD45RA+), Tfh (T follicular helper). Pre-vaccination (Pre), one week post two-dose COVID-19 mRNA vaccination (1w PV2), three months post two-dose vaccination (3m PV2), two weeks post third vaccination dose (2w PV3).

**Figure S5. Vaccination induces enhanced variant binding and neutralizing breadth in RBD-specific MBCs and antibody in PI as compared to N individuals.**

(A) Percent Naive B cells (CD21^+^CD27^-^) and (B) percent antigen-experienced (ag-exp.) B cells MBCs (CD21^+^CD27^+^ and CD21^-^CD27^+/-^) of RBD-specific non-plasmablasts in N and PI PBMCs at the indicated time points. (C) Percent IgG+ classical MBCs (CD21^+^CD27^+^) of RBD-specific ag-exp. B cells in N and PI PBMCs at the indicated time points. (D) Representative gating on RBD(Wu-1)-specific antigen-experienced (ag-exp.) B cells for RBD(Wu-1) and RBD(β) tetramer binding to identify cross-variant specific (Wu-1^+^β^+^) MBCs in N and PI PBMCs at the indicated time points. (E) Percent RBD(Wu-1^+^β^+^)-specific MBCs of RBD(Wu-1)-specific ag-exp. B cells in N and PI PBMCs at the indicated time points. (F) ELISA area under the curve (AUC) for RBD(β)-specific IgG plasma antibody from N and PI individuals at indicated time points. (G) Percent neutralization of SARS-CoV-2(Wu-1) virus by plasma from N and PI individuals at the indicated time points and (H) correlation with RBD(Wu-1)-specific IgG AUC. Statistics determined by two-tailed Mann-Whitney tests: not significant (ns), \**P* < 0.05, \*\**P* < 0.005, \*\*\**P* < 0.0005, and \*\*\*\**P* < 0.0001. Pre-vaccination (Pre), one week post two-dose COVID-19 mRNA vaccination (1w PV2), three months post two-dose vaccination (3m PV2), two weeks post third vaccination dose (2w PV3). Error bars represent mean and SD.

**Figure S6. Assessment of AIM CD4+ T helper memory subsets. Related to Figure 4.**

(**A**) Longitudinal summary graphs of indicated AIM CD4+ helper memory subsets for SARS-CoV-2 Naive (blue) and Previously Infected (red) donors. Significance was determined by Wilcoxon matched-paired signed rank test for longitudinal analyses and multiple unpaired Mann-Whitney test for group analyses: \**P* < 0.05, \*\**P* < 0.01, \*\*\**P* < 0.001, and \*\*\*\**P* < 0.0001. Error bars represent mean and SD. Dashed lines indicate average donor background level. Lines connecting data points indicate paired samples from the same donor. Pre-vaccination (Pre), one week post two-dose COVID-19 mRNA vaccination (1w PV2), three months post two-dose vaccination (3m PV2), two weeks post third vaccination dose (2w PV3).

**Figure S7. Gating Strategy and Backgating Validation of CD69^+^CD154^+^ AIM^+^ CD4+ T Cells. Related to Figure 4.**

(**A**) AIM^+^ gating strategy depicting removal of Naive CD45RA+CD127+ T Cells and CD25+CD127- T Regulatory Cells. (**B**) Validation of CD69+CD154+ AIM+ Cells depicting absence from T Regulatory and Naive compartments (left) and inclusion of all cytokine-producing cells (right).

**Figure S8. Assessment of Total CD69+CD154+ AIM+ CD4+ T Cells from *ex vivo* Cytokine Release Assay. Related to Figure 4.**

**(A)** Representative flow cytometry gating of non-naive (nn) CD69+CD154+ AIM+ CD4+ T Cells in DMSO-, M/N-, and S-stimulated memory T Cells from SARS-CoV-2 Naive (left, blue) and SARS-CoV-2 Previously Infected (right, red) donors before and one weeks after two doses of mRNA vaccine. (**B** and **C**) Summary graphs of MN- (B) and S-specific (C) nnCD4+ T Cells. Data in (B) and (C) are represented both longitudinally (left) and by cross-group comparisons (middle, right). Significance was determined by Wilcoxon matched-paired signed rank test for longitudinal analyses and multiple unpaired Mann-Whitney test for group analyses: \**P* < 0.05, \*\**P* < 0.01, \*\*\**P* < 0.001. Error bars represent mean and SD. Dashed lines indicate average donor background level. Pre-vaccination (Pre), one week post two-dose COVID-19 mRNA vaccination (1w PV2), three months post two-dose vaccination (3m PV2), two weeks post third vaccination dose (2w PV3).

**Figure S9. Supplemental Cytokine Production by S-specific AIM+ CD4+ T Cells. Related to Figure 4.**

(A thru H) Representative flow cytometry plots and summary graphs for non-naive CD4+CD69+CD154+ T cells (AIM CD4s) for the indicated cytokines. Data in (B) and (D) are represented both longitudinally (left) and by cross-group comparisons (middle, right). Validation of cytokine staining for cases of very low cytokine producers indicated in (E) thru (H) are provided using PMA and Ionomycin positive control (yellow). Significance was determined by Wilcoxon matched-paired signed rank test for longitudinal analyses and multiple unpaired Mann-Whitney test for group analyses: \**P* < 0.05, \*\**P* < 0.01, \*\*\**P* < 0.001. Error bars represent mean and SD. Dashed lines indicate average donor background level. Pre-vaccination (Pre), one week post two-dose COVID-19 mRNA vaccination (1w PV2), three months post two-dose vaccination (3m PV2), two weeks post third vaccination dose (2w PV3).

**Figure S10. Cytokine Production by M/N-Specific AIM+ CD4+ T Cells. Related to Figure 4.**

(A-E) Summary graphs of the indicated cytokines for M/N-specific CD4+CD69+CD154+ T cells (AIM+CD4+) in SARS-CoV-2 Naive (blue) and Previously Infected (red) donors. Data are represented both longitudinally (left) and by cross-group comparisons (middle, right). Significance was determined by Wilcoxon matched-paired signed rank test for longitudinal analyses and multiple unpaired Mann-Whitney test for group analyses: \**P* < 0.05, \*\**P* < 0.01, \*\*\**P* < 0.001. Error bars represent mean and SD. Dashed lines indicate average donor background level. Pre-vaccination (Pre), one week post two-dose COVID-19 mRNA vaccination (1w PV2), three months post two-dose vaccination (3m PV2), two weeks post third vaccination dose (2w PV3).

**Figure S11. UMAP and Clustering Analysis of SARS-CoV-2 S-Specific CD4+AIM+ T Cells. Related to Figure 4.**

(**A**) Dimensionality reduction and Phenograph-derived clustering (*k*=40) overlaid for total AIM+CD4+(CD154+CD69+) T Cells pooled from all donors (left). Heatmap expression of the indicated parameters over UMAP space (right). (**B-P**) All cells from the indicated clusters (CL) are represented in corresponding color from UMAP plot in (A) overlayed on all AIM+CD4+ T Cells and showing expression of select parameters (IL-2, IFN-γ, IL-10, CXCR5, CD127, CD25, IL-4, and IL-21). Enumeration of each cluster is per 1M non-naive (nn) CD4+ T Cells in SARS-CoV-2 Naive (blue) and SARS-CoV-2 Previously Infected (red) donors. Pre-vaccination (Pre), one week post two-dose COVID-19 mRNA vaccination (1w PV2), three months post two-dose vaccination (3m PV2). Significance was determined by multiple unpaired Mann-Whitney test for group analyses and by Wilcoxon matched-paired signed rank test for longitudinal analyses: \**P* < 0.05, \*\**P* < 0.01.

## Supplemental Tables

**Table S1:** Study Cohort

**Table S2:** SARS-CoV-2 Peptide Pool Sequences

**Table S3:** Key Resources

## References

Abu-Raddad, L.J., Chemaitelly, H., Ayoub, H.H., Yassine, H.M., Benslimane, F.M., Khatib, H.A.A., Tang, P., Hasan, M.R., Coyle, P., Kanaani, Z.A., et al. (2021). Protection afforded by the BNT162b2 and mRNA-1273 COVID-19 vaccines in fully vaccinated cohorts with and without prior infection. Medrxiv 2021.07.25.21261093.

Baden, L.R., Sahly, H.M.E., Essink, B., Kotloff, K., Frey, S., Novak, R., Diemert, D., Spector, S.A., Rouphael, N., Creech, C.B., et al. (2020). Efficacy and Safety of the mRNA-1273 SARS-CoV-2 Vaccine. New Engl J Medicine 384, NEJMoa2035389.

Becht, E., Tolstrup, D., Dutertre, C.-A., Morawski, P.A., Campbell, D.J., Ginhoux, F., Newell, E.W., Gottardo, R., and Headley, M.B. (2021). High-throughput single-cell quantification of hundreds of proteins using conventional flow cytometry and machine learning. Science Advances 7.

Brinkmann, V., Geiger, T., Alkan, S., Heusser, C.H. (1993) Interferon alpha increases the frequency of interferon gamma-producing human CD4+ T cells. J Exp Med. 178, 1655–63.

Cameroni, E., Bowen, J.E., Rosen, L.E., Saliba, C., Zepeda, S.K., Culap, K., Pinto, D., VanBlargan, L.A., Marco, A.D., Iulio, J. di, et al. (2021). Broadly neutralizing antibodies overcome SARS-CoV-2 Omicron antigenic shift. Nature 1–9.

Cancro, M.P., and Tomayko, M.M. (2021). Memory B cells and plasma cells: The differentiative continuum of humoral immunity. Immunol Rev 303, 72–82.

Cho, A., Muecksch, F., Schaefer-Babajew, D., Wang, Z., Finkin, S., Gaebler, C., Ramos, V., Cipolla, M., Mendoza, P., Agudelo, M., et al. (2021). Anti-SARS-CoV-2 receptor-binding domain antibody evolution after mRNA vaccination. Nature 600, 517–522.

Collier, D.A., Marco, A.D., Ferreira, I.A.T.M., Meng, B., Datir, R.P., Walls, A.C., Kemp, S.A., Bassi, J., Pinto, D., Silacci-Fregni, C., et al. (2021). Sensitivity of SARS-CoV-2 B.1.1.7 to mRNA vaccine-elicited antibodies. Nature 593, 136–141.

Corbett, K.S., Nason, M.C., Flach, B., Gagne, M., O’Connell, S., Johnston, T.S., Shah, S.N., Edara, V.V., Floyd, K., Lai, L., et al. (2021). Immune correlates of protection by mRNA-1273 vaccine against SARS-CoV-2 in nonhuman primates. Science 373, eabj0299.

Crawford, K.H.D., Eguia, R., Dingens, A.S., Loes, A.N., Malone, K.D., Wolf, C.R., Chu, H.Y., M, A.T., Veesler, D., Murphy, M., et al. (2020). Protocol and Reagents for Pseudotyping Lentiviral Particles with SARS-CoV-2 Spike Protein for Neutralization Assays. Viruses 12, 513.

Crotty, S. (2021). Hybrid immunity. Science 372, 1392–1393.

Dan, J.M., Mateus, J., Kato, Y., Hastie, K.M., Yu, E.D., Faliti, C.E., Grifoni, A., Ramirez, S.I., Haupt, S., Frazier, A., et al. (2021). Immunological memory to SARS-CoV-2 assessed for up to 8 months after infection. Science 371, eabf4063.

De Waal Malefyt, R., Abrams, J., Bennett, B., Figdor C.G., de Vries J.E. (1991) Interleukin 10 (IL-10) Inhibits Cytokine Synthesis by Human Monocytes: An Autoregulatory Role of IL-10 Produced by Monocytes. J. Exp. Med. 174, 1209–1220.

Erasmus, J.H., Khandhar, A.P., O’Connor, M.A., Walls, A.C., Hemann, E.A., Murapa, P., Archer, J., Leventhal, S., Fuller, J.T., Lewis, T.B., et al. (2020). An Alphavirus-derived replicon RNA vaccine induces SARS-CoV-2 neutralizing antibody and T cell responses in mice and nonhuman primates. Sci Transl Med 12, eabc9396.

Feng, S., Phillips, D.J., White, T., Sayal, H., Aley, P.K., Bibi, S., Dold, C., Fuskova, M., Gilbert, S.C., Hirsch, I., et al. (2021). Correlates of protection against symptomatic and asymptomatic SARS-CoV-2 infection. Nat Med 27, 2032–2040.

Gabryšová, L., Alvarez-Martinez, M., Luisier, R., Cox, L.S., Sodenkamp, J., Hosking, C., Pérez-Mazliah, D., Whicher, C., Kannan, Y., Potempa, K., et al. (2018). c-Maf controls immune responses by regulating disease-specific gene networks and repressing IL-2 in CD4+ T cells. Nat Immunol 19, 497–507.

Galani, I.-E., Rovina, N., Lampropoulou, V., Triantafyllia, V., Manioudaki, M., Pavlos, E., Koukaki, E., Fragkou, P.C., Panou, V., Rapti, V., et al. (2021). Untuned antiviral immunity in COVID-19 revealed by temporal type I/III interferon patterns and flu comparison. Nat Immunol 22, 32–40.

Gazit, S., Shlezinger, R., Perez, G., Lotan, R., Peretz, A., Ben-Tov, A., Cohen, D., Muhsen, K., Chodick, G., and Patalon, T. (2021). Comparing SARS-CoV-2 natural immunity to vaccine-induced immunity: reinfections versus breakthrough infections. Medrxiv 2021.08.24.21262415.

Gazzinelli, R.T., Wysocka, M., Hieny, S., Scharton-Kersten, T., Cheever, A., Kühn R., Müller, W., Trinchieri, G., and Sher, A. (1996) In the absence of endogenous IL-10, mice acutely infected with Toxoplasma gondii succumb to a lethal immune response dependent on CD4+ T cells and accompanied by overproduction of IL-12, IFN-gamma and TNF-alpha. J. Immunology 157, 798–805.

Gilbert, P.B., Montefiori, D.C., McDermott, A.B., Fong, Y., Benkeser, D., Deng, W., Zhou, H., Houchens, C.R., Martins, K., Jayashankar, L., et al. (2021). Immune correlates analysis of the mRNA-1273 COVID-19 vaccine efficacy clinical trial. Science 375, 43–50.

Glass, D.R., Tsai, A.G., Oliveria, J.P., Hartmann, F.J., Kimmey, S.C., Calderon, A.A., Borges, L., Glass, M.C., Wagar, L.E., Davis, M.M., et al. (2020). An Integrated Multi-omic Single-Cell Atlas of Human B Cell Identity. Immunity 53, 217–232.e5.

Goel, R.R., Painter, M.M., Apostolidis, S.A., Mathew, D., Meng, W., Rosenfeld, A.M., Lundgreen, K.A., Reynaldi, A., Khoury, D.S., Pattekar, A., et al. (2021). mRNA vaccines induce durable immune memory to SARS-CoV-2 and variants of concern. Science eabm0829.

Goldberg, Y., Mandel, M., Bar-On, Y.M., Bodenheimer, O., Freedman, L., Ash, N., Alroy-Preis, S., Huppert, A., and Milo, R. (2021). Protection and waning of natural and hybrid COVID-19 immunity. Medrxiv 2021.12.04.21267114.

Guerrera, G., Picozza, M., D’Orso, S., Placido, R., Pirronello, M., Verdiani, A., Termine, A., Fabrizio, C., Giannessi, F., Sambucci, M., et al. (2021). BNT162b2 vaccination induces durable SARS-CoV-2 specific T cells with a stem cell memory phenotype. Sci Immunol 6, eabl5344.

Hunter, C.A., Ellis-Neyes, L.A., Slifer, T., Kanaly, S., Griinig, G., Fort, M., Rennick, D., and Araujot, F.G. (1997) 1L-10 Is Required to Prevent Immune Hyperactivity During Infection with Trypanosoma cruzi. J. Immunology 158, 3311–3316.

Jankovic, D., Kullberg, M.C., Feng, C.G., Goldszmid, R.S., Collazo, C.M., Wilson, M., Wynn, T.A., Kamanaka, M., Flavell, R.A., and Sher, A. (2007). Conventional T-bet+Foxp3− Th1 cells are the major source of host-protective regulatory IL-10 during intracellular protozoan infection. J Exp Medicine 204, 273–283.

Khoury, D.S., Cromer, D., Reynaldi, A., Schlub, T.E., Wheatley, A.K., Juno, J.A., Subbarao, K., Kent, S.J., Triccas, J.A., and Davenport, M.P. (2021). Neutralizing antibody levels are highly predictive of immune protection from symptomatic SARS-CoV-2 infection. Nat Med 27, 1205–1211.

Kim, C.C., Baccarella, A.M., Bayat, A., Pepper, M., and Fontana, M.F. (2019). FCRL5+ Memory B Cells Exhibit Robust Recall Responses. Cell Reports 27, 1446–1460.e4.

Kim, W., Zhou, J.Q., Sturtz, A.J., Horvath, S.C., Schmitz, A.J., Lei, T., Kalaidina, E., Thapa, M., Alsoussi, W.B., Haile, A., et al. (2021). Germinal centre-driven maturation of B cell response to SARS-CoV-2 vaccination. Biorxiv 2021.10.31.466651.

Klicznik, M.M., Morawski, P.A., Höllbacher, B., Varkhande, S.R., Motley, S.J., Kuri-Cervantes, L., Goodwin, E., Rosenblum, M.D., Long, S.A., Brachtl, G., et al. (2019). Human CD4+CD103+ cutaneous resident memory T cells are found in the circulation of healthy individuals. Sci Immunol 4, eaav8995.

Knox, J.J., Kaplan, D.E., and Betts, M.R. (2017). T-bet-expressing B cells during HIV and HCV infections. Cell Immunol 321, 26–34.

Krishnamurty, A.T., Thouvenel, C.D., Portugal, S., Keitany, G.J., Kim, K.S., Holder, A., Crompton, P.D., Rawlings, D.J., and Pepper, M. (2016). Somatically Hypermutated Plasmodium-Specific IgM+ Memory B Cells Are Rapid, Plastic, Early Responders upon Malaria Rechallenge. Immunity 45, 402–414.

Laidlaw, B.J., and Ellebedy, A.H. (2021). The germinal centre B cell response to SARS-CoV-2. Nat Rev Immunol 22, 1–12.

Lazarevic, V., Glimcher, L.H., and Lord, G.M. (2013). T-bet: a bridge between innate and adaptive immunity. Nat Rev Immunol 13, 777–789.

Lei, X., Dong, X., Ma, R., Wang, W., Xiao, X., Tian, Z., Wang, C., Wang, Y., Li, L., Ren, L., et al. (2020). Activation and evasion of type I interferon responses by SARS-CoV-2. Nat Commun 11, 3810.

Levine, J.H., Simonds, E.F., Bendall, S.C., Davis, K.L., Amir, E.D., Tadmor, M.D., Litvin, O., Fienberg, H.G., Jager, A., Zunder, E.R., et al. (2015). Data-Driven Phenotypic Dissection of AML Reveals Progenitor-like Cells that Correlate with Prognosis. Cell 162, 184–197.

Liu, G., Lee, J.-H., Parker, Z.M., Acharya, D., Chiang, J.J., Gent, M. van, Riedl, W., Davis-Gardner, M.E., Wies, E., Chiang, C., et al. (2021). ISG15-dependent activation of the sensor MDA5 is antagonized by the SARS-CoV-2 papain-like protease to evade host innate immunity. Nat Microbiol 6, 467–478.

Liu, Y., Qin, C., Rao, Y., Ngo, C., Feng, J.J., Zhao, J., Zhang, S., Wang, T.-Y., Carriere, J., Savas, A.C., et al. (2021). SARS-CoV-2 Nsp5 Demonstrates Two Distinct Mechanisms Targeting RIG-I and MAVS To Evade the Innate Immune Response. Mbio 12, e02335–21.

Lucas, C., Wong, P., Klein, J., Castro, T.B., Silva, J., Sundaram, M., Ellingson, M.K., Mao, T., Oh, J., Israelow, B., et al. (2020). Longitudinal analyses reveal immunological misfiring in severe COVID-19. Nature 1–9.

O’Garra, A., and Vieira, P. (2007). TH1 cells control themselves by producing interleukin-10. Nat Rev Immunol 7, 425–428.

Olatunde, A.C., Hale, J.S., and Lamb, T.J. (2021). Cytokine-skewed Tfh cells: functional consequences for B cell help. Trends Immunol 42, 536–550.

Painter, M.M., Mathew, D., Goel, R.R., Apostolidis, S.A., Pattekar, A., Kuthuru, O., Baxter, A.E., Herati, R.S., Oldridge, D.A., Gouma, S., et al. (2021). Rapid induction of antigen-specific CD4+ T cells is associated with coordinated humoral and cellular immunity to SARS-CoV-2 mRNA vaccination. Immunity 54, 2133–2142.e3.

Polack, F.P., Thomas, S.J., Kitchin, N., Absalon, J., Gurtman, A., Lockhart, S., Perez, J.L., Marc, G.P., Moreira, E.D., Zerbini, C., et al. (2020). Safety and Efficacy of the BNT162b2 mRNA Covid-19 Vaccine. New Engl J Medicine 383, NEJMoa2034577.

Poon, M.M.L., Rybkina, K., Kato, Y., Kubota, M., Matsumoto, R., Bloom, N.I., Zhang, Z., Hastie, K.M., Grifoni, A., Weiskopf, D., et al. (2021). SARS-CoV-2 infection generates tissue-localized immunological memory in humans. Sci Immunol 6, eabl9105.

Pot, C., Jin, H., Awasthi, A., Liu, S.M., Lai, C.-Y., Madan, R., Sharpe, A.H., Karp, C.L., Miaw, S.-C., Ho, I.-C., et al. (2009). Cutting Edge: IL-27 Induces the Transcription Factor c-Maf, Cytokine IL-21, and the Costimulatory Receptor ICOS that Coordinately Act Together to Promote Differentiation of IL-10-Producing Tr1 Cells. J Immunol 183, 797–801.

Reiss, S., Baxter, A.E., Cirelli, K.M., Dan, J.M., Morou, A., Daigneault, A., Brassard, N., Silvestri, G., Routy, J.-P., Havenar-Daughton, C., et al. (2017). Comparative analysis of activation induced marker (AIM) assays for sensitive identification of antigen-specific CD4 T cells. Plos One 12, e0186998.

Reynolds, C.J., Pade, C., Gibbons, J.M., Butler, D.K., Otter, A.D., Menacho, K., Fontana, M., Smit, A., Sackville-West, J.E., Cutino-Moguel, T., et al. (2021). Prior SARS-CoV-2 infection rescues B and T cell responses to variants after first vaccine dose. Science 372, 1418–1423.

Rodda, L.B., Netland, J., Shehata, L., Pruner, K.B., Morawski, P.A., Thouvenel, C.D., Takehara, K.K., Eggenberger, J., Hemann, E.A., Waterman, H.R., et al. (2021). Functional SARS-CoV-2-Specific Immune Memory Persists after Mild COVID-19. Cell 184, 169–183.e17.

Ruterbusch, M., Pruner, K.B., Shehata, L., and Pepper, M. (2020). In Vivo CD4+ T Cell Differentiation and Function: Revisiting the Th1/Th2 Paradigm. Annu Rev Immunol 38, 705–725.

Schmidt, F., Muecksch, F., Weisblum, Y., Da Silva, J., Bednarski, E., Cho, A., Wang, Z., Gaebler, C., Caskey, M., Nussenzweig, M.C., et al. (2021). Plasma Neutralization of the SARS-CoV-2 Omicron Variant. New England Journal of Medicine.

Schmidt, F., Weisblum, Y., Rutkowska, M., Poston, D., DaSilva, J., Zhang, F., Bednarski, E., Cho, A., Schaefer-Babajew, D.J., Gaebler, C., et al. (2021). High genetic barrier to SARS-CoV-2 polyclonal neutralizing antibody escape. Nature 600, 512–516.

Stamatatos, L., Czartoski, J., Wan, Y.-H., Homad, L.J., Rubin, V., Glantz, H., Neradilek, M., Seydoux, E., Jennewein, M.F., MacCamy, A.J., et al. (2021). mRNA vaccination boosts cross-variant neutralizing antibodies elicited by SARS-CoV-2 infection. Sci New York N Y 372, eabg9175.

Sun, J., Madan, R., Karp, C.L., and Braciale, T.J. (2009). Effector T cells control lung inflammation during acute influenza virus infection by producing IL-10. Nat Med 15, 277–284.

Tarke, A., Sidney, J., Kidd, C.K., Dan, J.M., Ramirez, S.I., Yu, E.D., Mateus, J., Antunes, R. da S., Moore, E., Rubiro, P., et al. (2021). Comprehensive analysis of T cell immunodominance and immunoprevalence of SARS-CoV-2 epitopes in COVID-19 cases. Cell Reports Medicine 2, 100204.

Thoms, M., Buschauer, R., Ameismeier, M., Koepke, L., Denk, T., Hirschenberger, M., Kratzat, H., Hayn, M., Mackens-Kiani, T., Cheng, J., et al. (2020). Structural basis for translational shutdown and immune evasion by the Nsp1 protein of SARS-CoV-2. Sci New York N Y 369, 1249–1255.

Tortorici, M.A., Czudnochowski, N., Starr, T.N., Marzi, R., Walls, A.C., Zatta, F., Bowen, J.E., Jaconi, S., Iulio, J.D., Wang, Z., et al. (2021). Broad sarbecovirus neutralization by a human monoclonal antibody. Nature 597, 103–108.

Turner, J.S., O’Halloran, J.A., Kalaidina, E., Kim, W., Schmitz, A.J., Zhou, J.Q., Lei, T., Thapa, M., Chen, R.E., Case, J.B., et al. (2021). SARS-CoV-2 mRNA vaccines induce persistent human germinal centre responses. Nature 596, 109–113.

Walls, A.C., Fiala, B., Schäfer, A., Wrenn, S., Pham, M.N., Murphy, M., Tse, L.V., Shehata, L., O’Connor, M.A., Chen, C., et al. (2020). Elicitation of Potent Neutralizing Antibody Responses by Designed Protein Nanoparticle Vaccines for SARS-CoV-2. Cell 183, 1367–1382.e17.

Wang, Z., Muecksch, F., Schaefer-Babajew, D., Finkin, S., Viant, C., Gaebler, C., Hoffmann, H.-H., Barnes, C.O., Cipolla, M., Ramos, V., et al. (2021). Naturally enhanced neutralizing breadth against SARS-CoV-2 one year after infection. Nature 595, 426–431.

Wijeyesinghe, S., Beura, L.K., Pierson, M.J., Stolley, J.M., Adam, O.A., Ruscher, R., Steinert, E.M., Rosato, P.C., Vezys, V., and Masopust, D. (2021). Expansible residence decentralizes immune homeostasis. Nature 1–6.

Xia, H., Cao, Z., Xie, X., Zhang, X., Chen, J.Y.-C., Wang, H., Menachery, V.D., Rajsbaum, R., and Shi, P.-Y. (2020). Evasion of Type I Interferon by SARS-CoV-2. Cell Reports 33, 108234–108234.

Yang, Y., Ochando, J., Yopp, A., Bromberg, J.S., and Ding, Y. (2005). IL-6 Plays a Unique Role in Initiating c-Maf Expression during Early Stage of CD4 T Cell Activation. J Immunol 174, 2720–2729.

Zhao, J., Zhao, J., Mangalam, A.K., Channappanavar, R., Fett, C., Meyerholz, D.K., Agnihothram, S., Baric, R.S., David, C.S., and Perlman, S. (2016). Airway Memory CD4+ T Cells Mediate Protective Immunity against Emerging Respiratory Coronaviruses. Immunity 44, 1379–1391.

