## Supplemental Figures and Tables for "Imprinted SARS-CoV-2-specific memory lymphocytes define hybrid immunity"

Figure S1

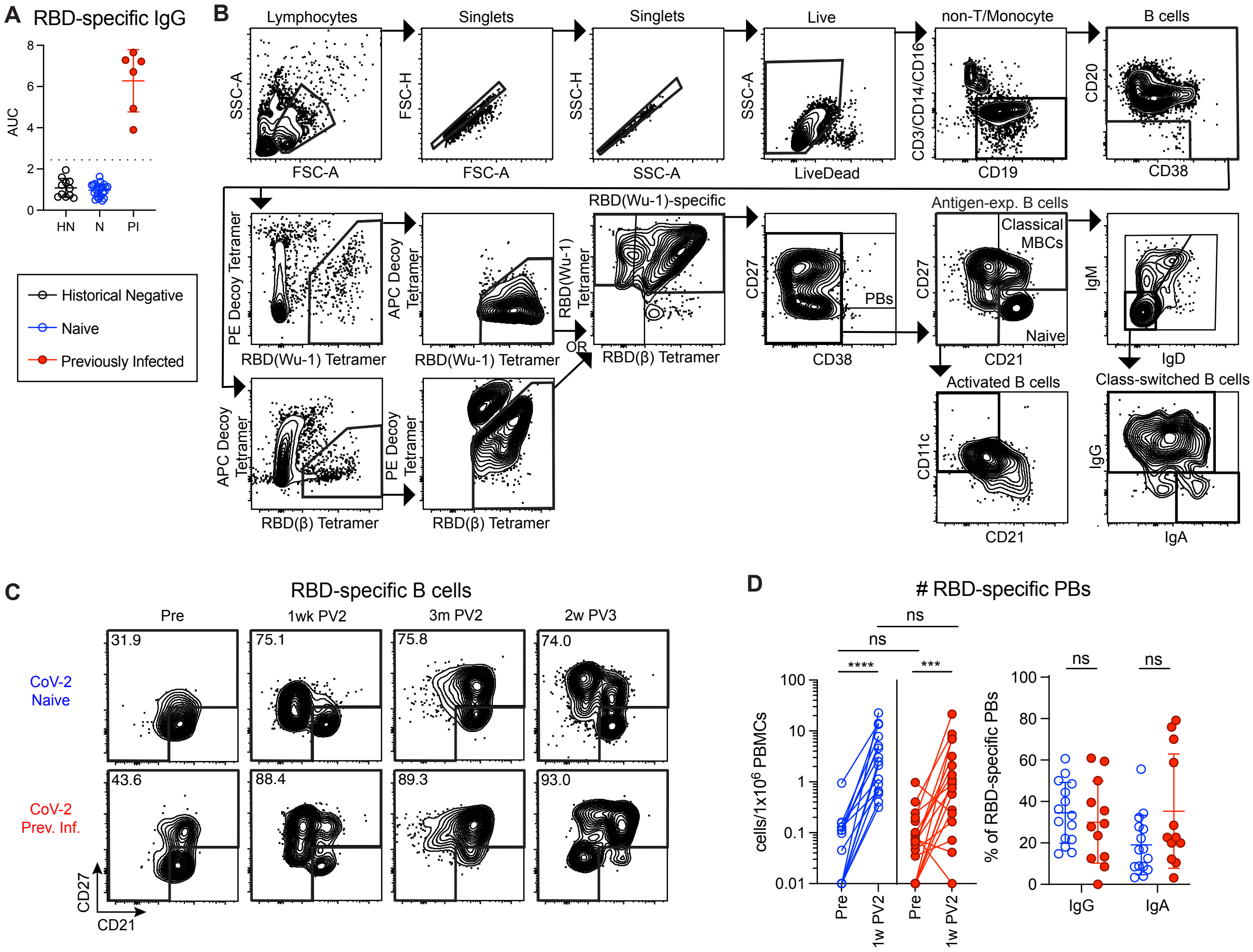

A RBD-specific IgG
AUC
HN
N
PI
Historical Negative
Naive
Previously Infected
B
Lymphocytes
Singlets
Singlets
Live
non-T/Monocyte
B cells
SSC-A
FSC-A
FSC-H
FSC-A
SSC-H
SSC-A
SSC-A
LiveDead
CD3/CD14/CD16
CD19
CD20
CD38
PE Decoy Tetramer
RBD(Wu-1) Tetramer
APC Decoy Tetramer
RBD(Wu-1) Tetramer
RBD(Wu-1)-specific
RBD(Wu-1) Tetramer
RBD(β) Tetramer
OR
CD27
CD38
PBs
Antigen-exp. B cells
Classical MBCs
Naive
CD27
CD21
IgM
IgD
APC Decoy Tetramer
RBD(β) Tetramer
PE Decoy Tetramer
RBD(β) Tetramer
Activated B cells
CD11c
CD21
Class-switched B cells
IgG
IgA
C
RBD-specific B cells
Pre
1wk PV2
3m PV2
2w PV3
CoV-2 Naive
31.9
75.1
75.8
74.0
CoV-2 Prev. Inf.
43.6
88.4
89.3
93.0
CD27
CD21
D
### RBD-specific PBs
cells/1x10^6 PBMCs
Pre
1w PV2
Pre
1w PV2
ns
ns
****
***
% of RBD-specific PBs
ns
ns
IgG
IgA

Figure S2

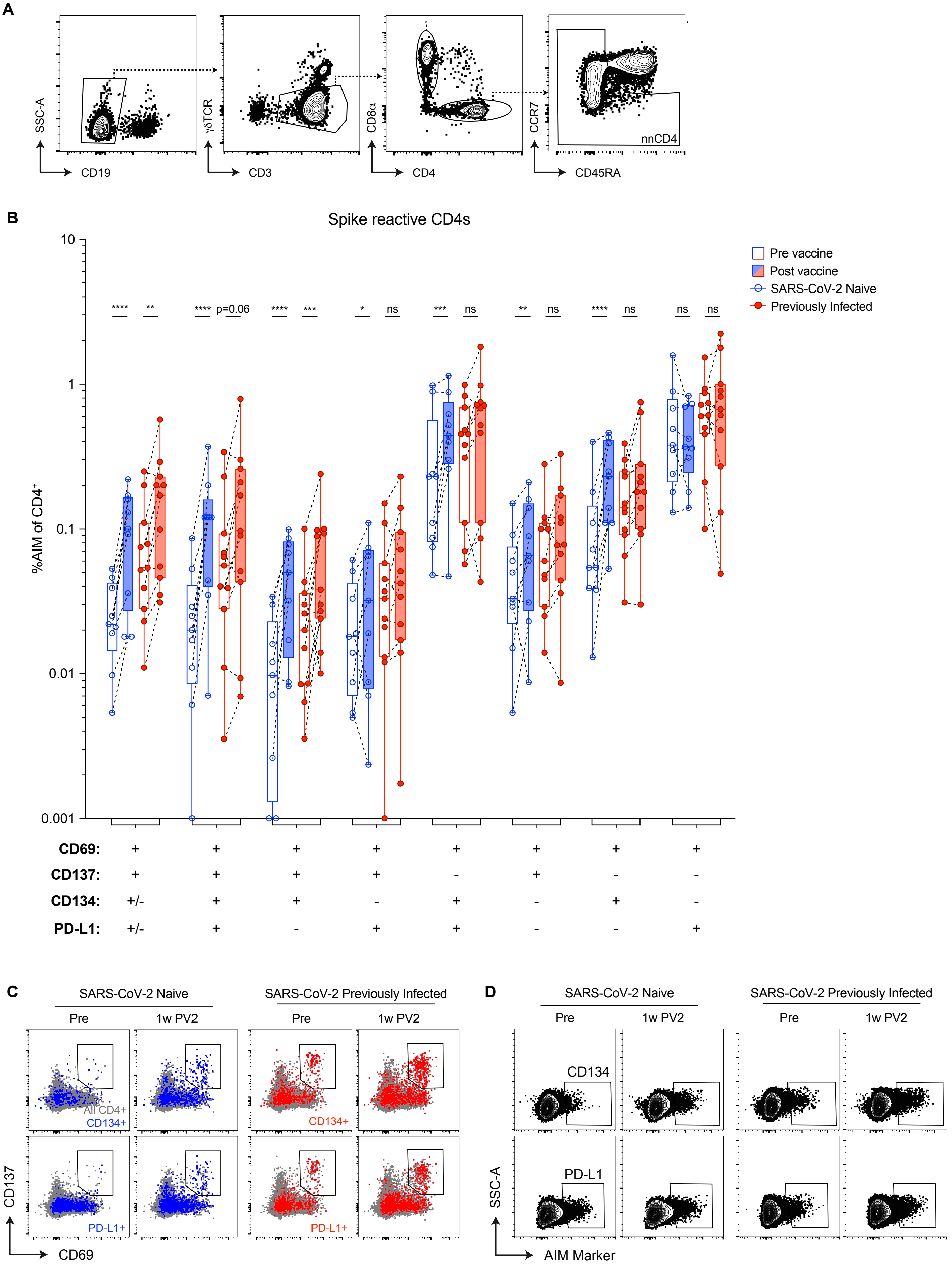
A
SSC-A
CD19
γδTCR
CD3
CD8α
CD4
CCR7
CD45RA
nnCD4
B
Spike reactive CD4s
%AIM of CD4+
10
1
0.1
0.01
0.001
Pre vaccine
Post vaccine
SARS-CoV-2 Naive
Previously Infected
**** ** **** p=0.06 **** *** * ns *** ns ** ns **** ns ns ns
CD69: + + + + + + + +
CD137: + + + + - + - -
CD134: +/- + + - + - + -
PD-L1: +/- + - + + - - +
C
SARS-CoV-2 Naive
SARS-CoV-2 Previously Infected
Pre
1w PV2
All CD4+
CD134+
PD-L1+
CD137
CD69
D
SARS-CoV-2 Naive
SARS-CoV-2 Previously Infected
Pre
1w PV2
CD134
PD-L1
SSC-A
AIM Marker

Figure S3

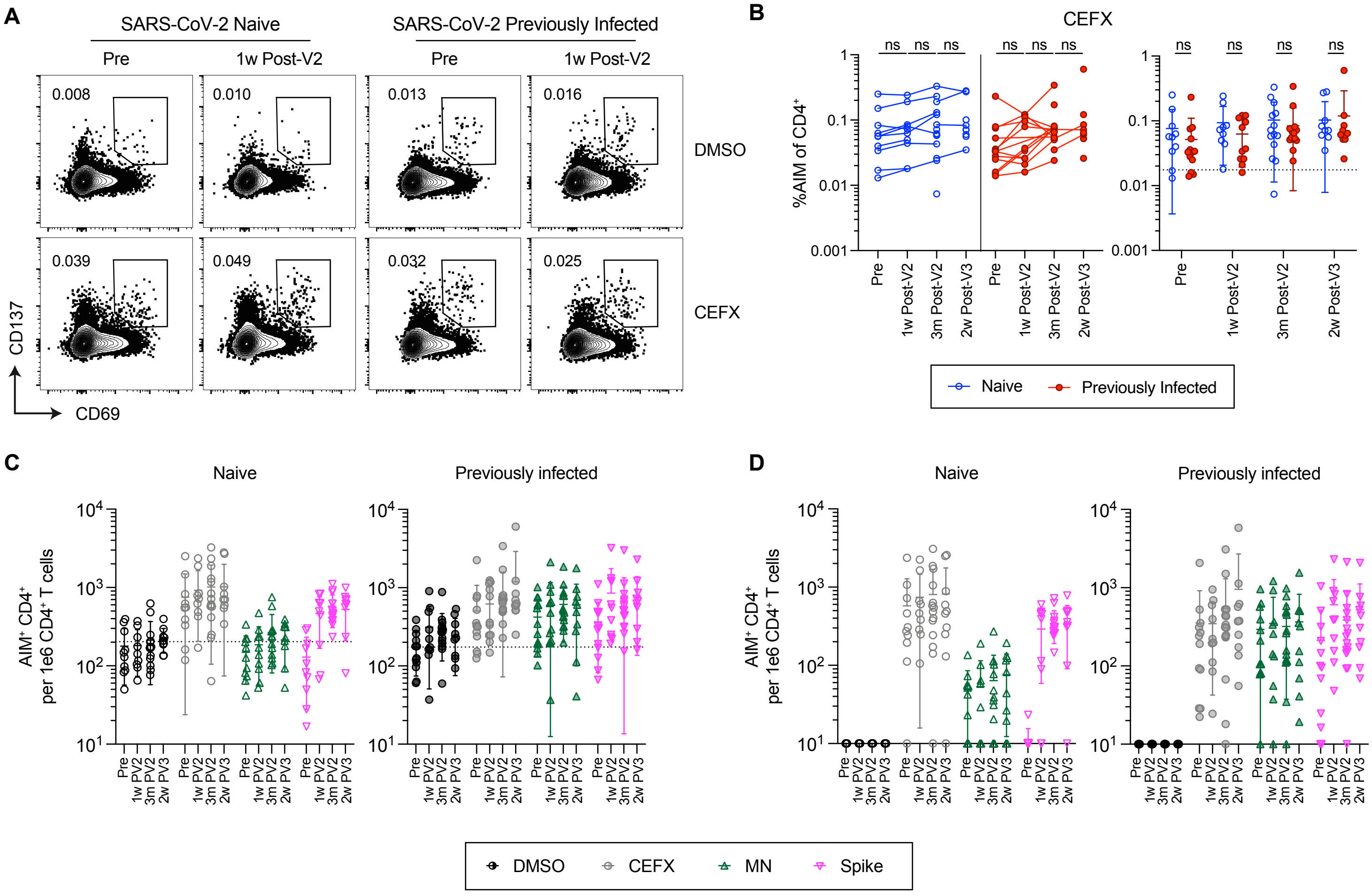
A
SARS-CoV-2 Naive
SARS-CoV-2 Previously Infected
Pre
1w Post-V2
0.008
0.010
0.013
0.016
DMSO
0.039
0.049
0.032
0.025
CEFX
CD137
CD69
B
CEFX
%AIM of CD4+
ns
Pre
1w Post-V2
3m Post-V2
2w Post-V3
Naive
Previously Infected
C
Naive
Previously infected
AIM+ CD4+ per 1e6 CD4+ T cells
Pre
1w PV2
3m PV2
2w PV3
D
Naive
Previously infected
DMSO
CEFX
MN
Spike

Figure S4

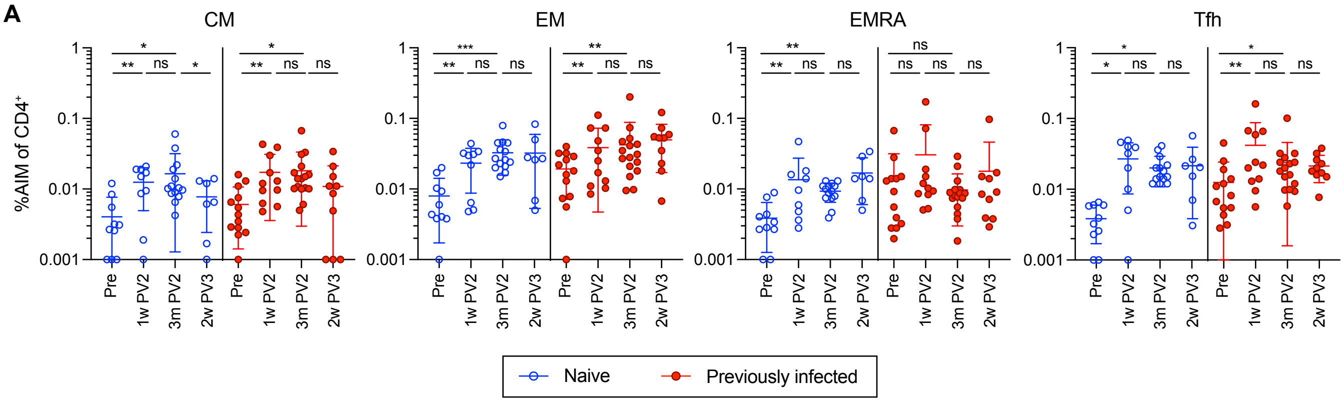
A
CM
EM
EMRA
Tfh
%AIM of CD4+
1
0.1
0.01
0.001
Pre
1w PV2
3m PV2
2w PV3
*
**
***
ns
Naive
Previously infected

#### Figure S5

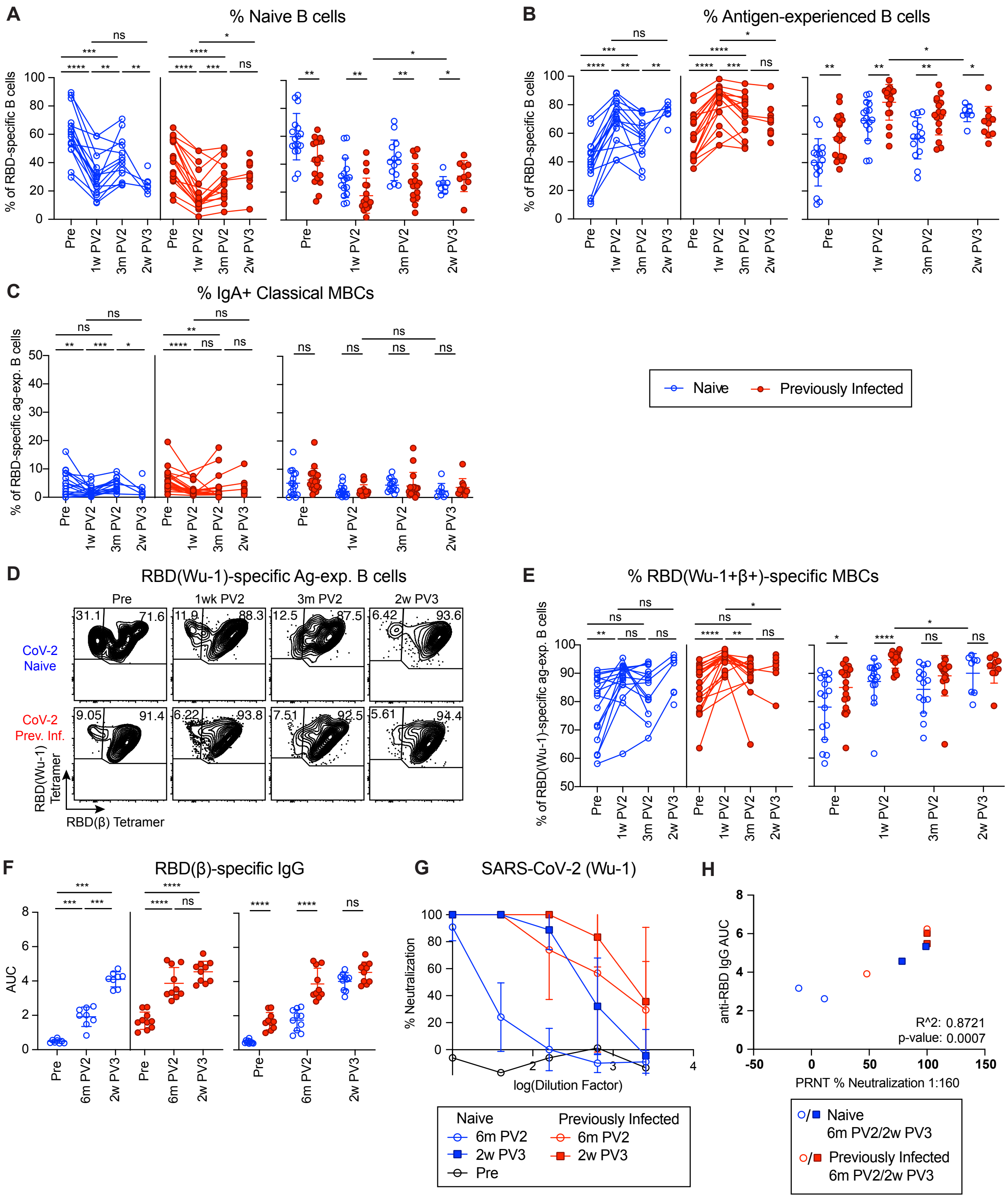

A
% Naive B cells
% of RBD-specific B cells
Pre
1w PV2
3m PV2
2w PV3
B
% Antigen-experienced B cells
C
% IgA+ Classical MBCs
% of RBD-specific ag-exp. B cells
Naive
Previously Infected
D
RBD(Wu-1)-specific Ag-exp. B cells
Pre
1wk PV2
3m PV2
2w PV3
CoV-2 Naive
CoV-2 Prev. Inf.
RBD(Wu-1) Tetramer
RBD(β) Tetramer
E
% RBD(Wu-1+β+)-specific MBCs
% of RBD(Wu-1)-specific ag-exp. B cells
F
RBD(β)-specific IgG
AUC
6m PV2
G
SARS-CoV-2 (Wu-1)
% Neutralization
log(Dilution Factor)
Naive
6m PV2
2w PV3
Pre
Previously Infected
6m PV2
2w PV3
H
anti-RBD IgG AUC
PRNT % Neutralization 1:160
R^2: 0.8721
p-value: 0.0007
Naive
6m PV2/2w PV3
Previously Infected
6m PV2/2w PV3

Figure S6

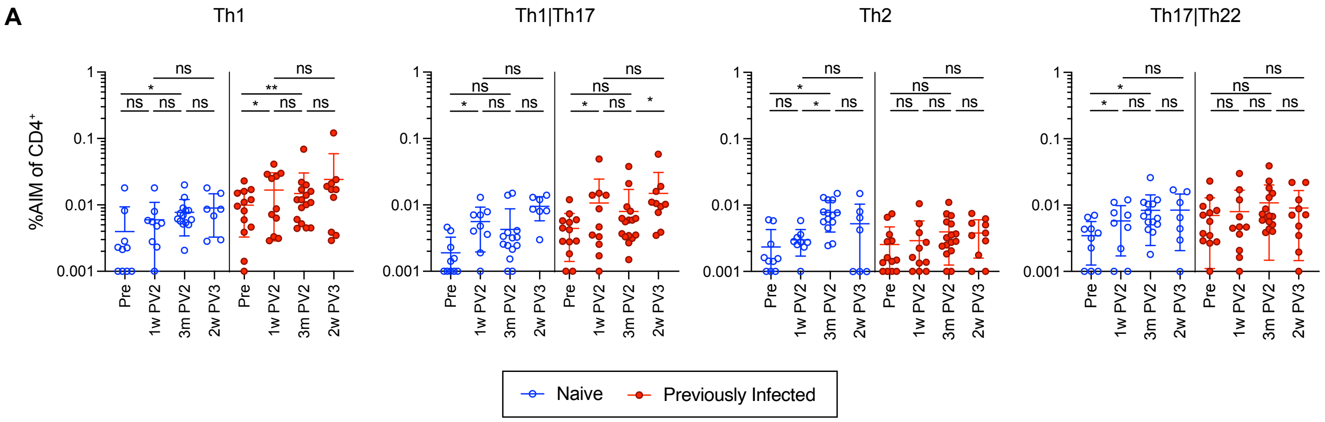

A
Th1
Th1|Th17
Th2
Th17|Th22
%AIM of CD4+
1
0.1
0.01
0.001
ns
*
**
Pre
1w PV2
3m PV2
2w PV3
Naive
Previously Infected

Figure S7

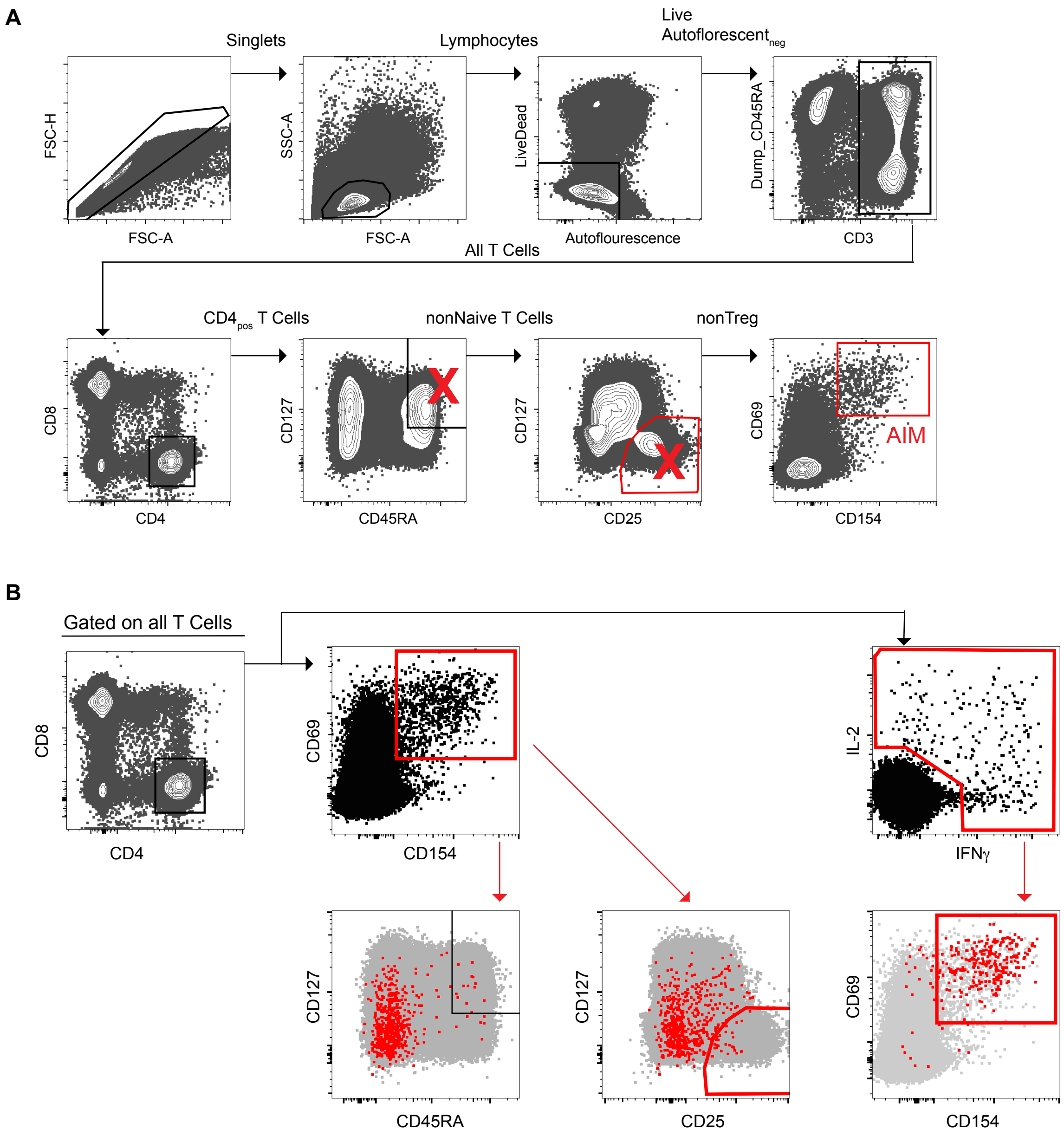
A
Singlets
Lymphocytes
Live
Autoflorescent neg
FSC-H
FSC-A
SSC-A
FSC-A
LiveDead
Autoflourescence
Dump_CD45RA
CD3
All T Cells
CD4 pos T Cells
nonNaive T Cells
nonTreg
CD8
CD4
CD127
CD45RA
CD127
CD25
CD69
CD154
AIM
B
Gated on all T Cells
CD8
CD4
CD69
CD154
IL-2
IFNγ
CD127
CD45RA
CD127
CD25
CD69
CD154

### Figure S8

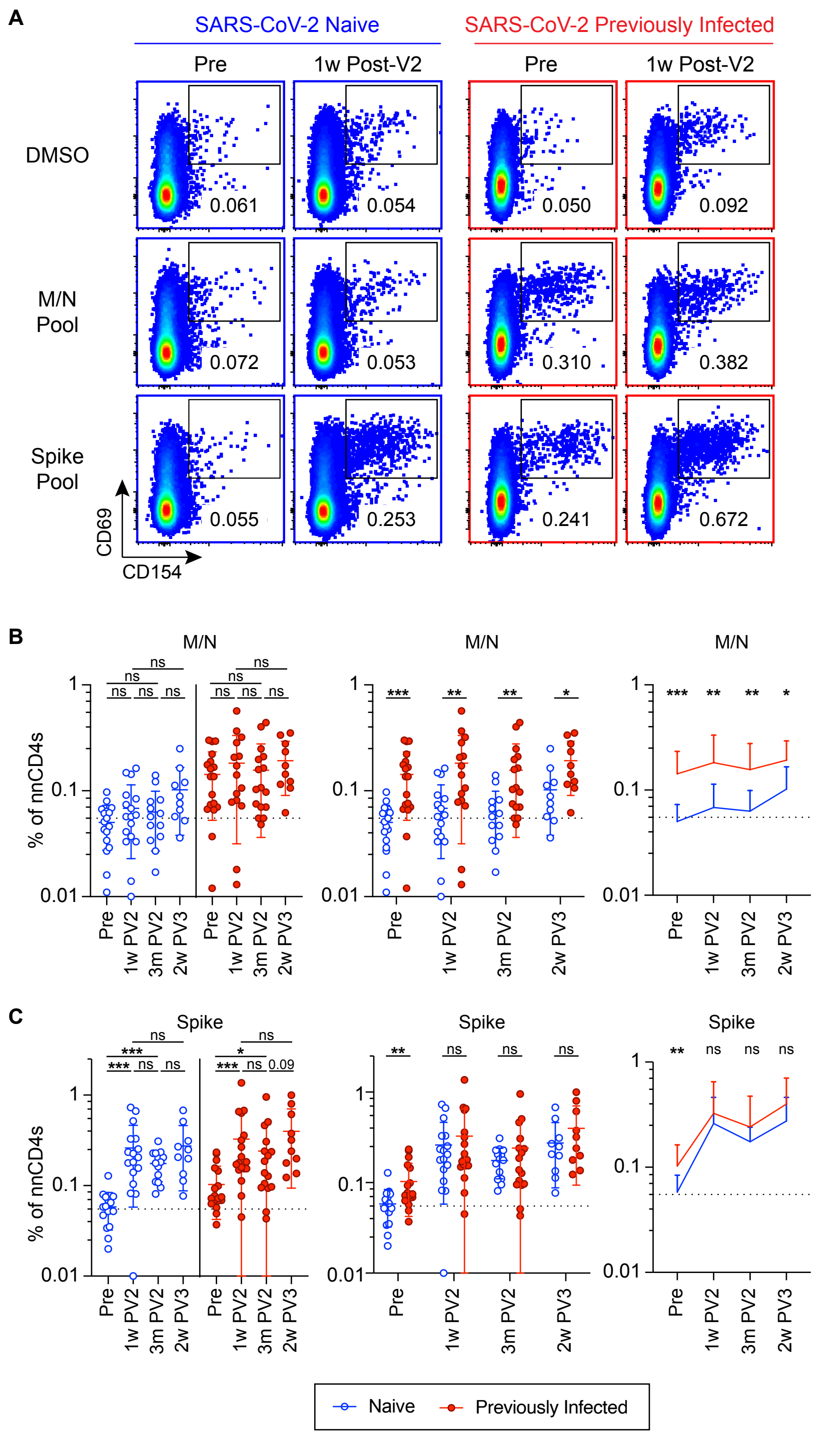
A
SARS-CoV-2 Naive
SARS-CoV-2 Previously Infected
Pre
1w Post-V2
Pre
1w Post-V2
DMSO
M/N Pool
Spike Pool
0.061
0.054
0.050
0.092
0.072
0.053
0.310
0.382
0.055
0.253
0.241
0.672
CD69
CD154
B
M/N
% of nnCD4s
Pre
1w PV2
3m PV2
2w PV3
ns
***
**
*
C
Spike
0.09
Naive
Previously Infected

Figure S9

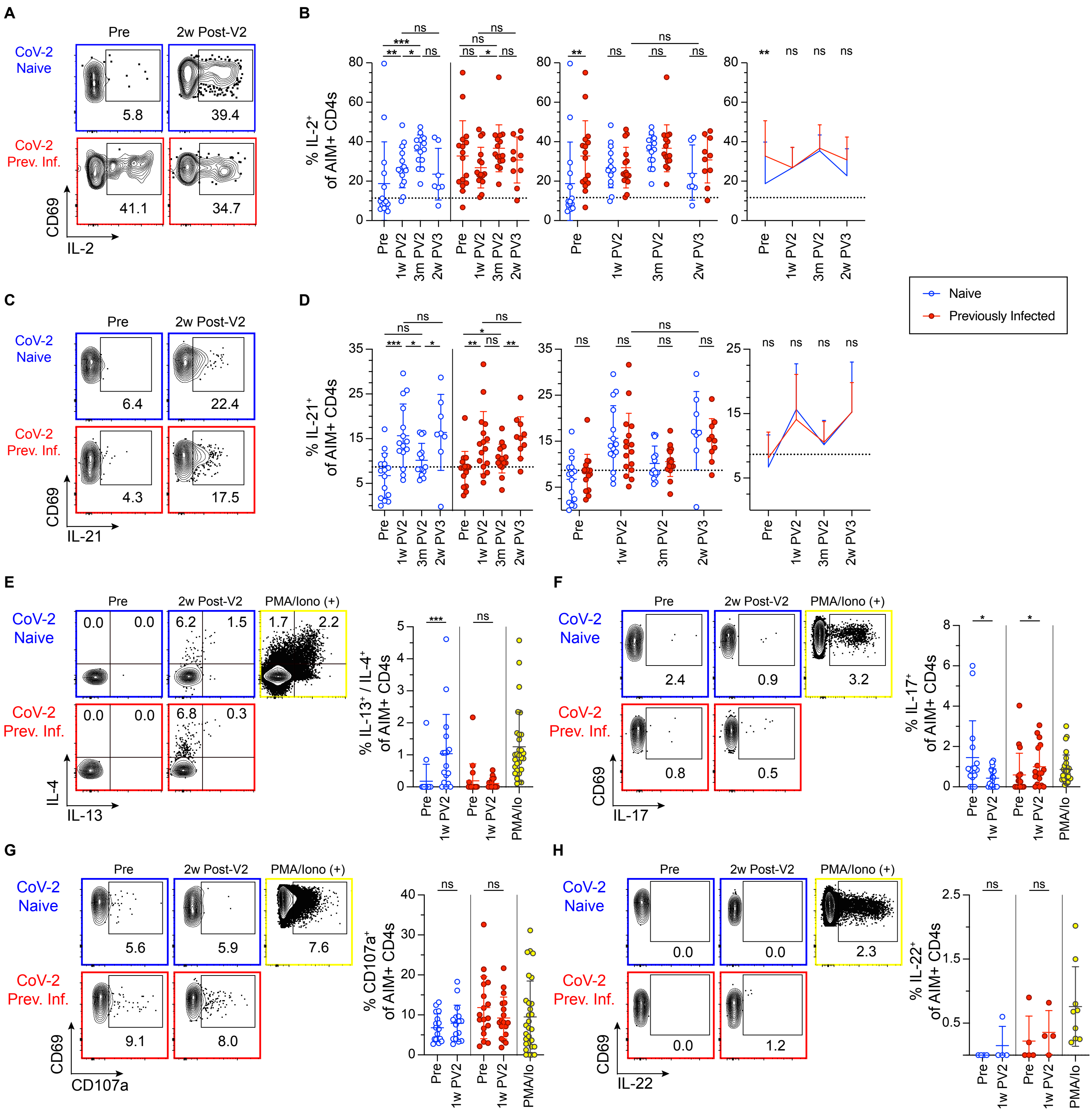
A
B
C
D
E
F
G
H
Pre
2w Post-V2
PMA/Iono (+)
CoV-2 Naive
CoV-2 Prev. Inf.
CD69
IL-2
IL-21
IL-4
IL-13
IL-17
CD107a
IL-22
% IL-2+ of AIM+ CD4s
% IL-21+ of AIM+ CD4s
% IL-13+ / IL-4+ of AIM+ CD4s
% IL-17+ of AIM+ CD4s
% CD107a+ of AIM+ CD4s
% IL-22+ of AIM+ CD4s
1w PV2
3m PV2
2w PV3
PMA/Io
Naive
Previously Infected
ns

Figure S10

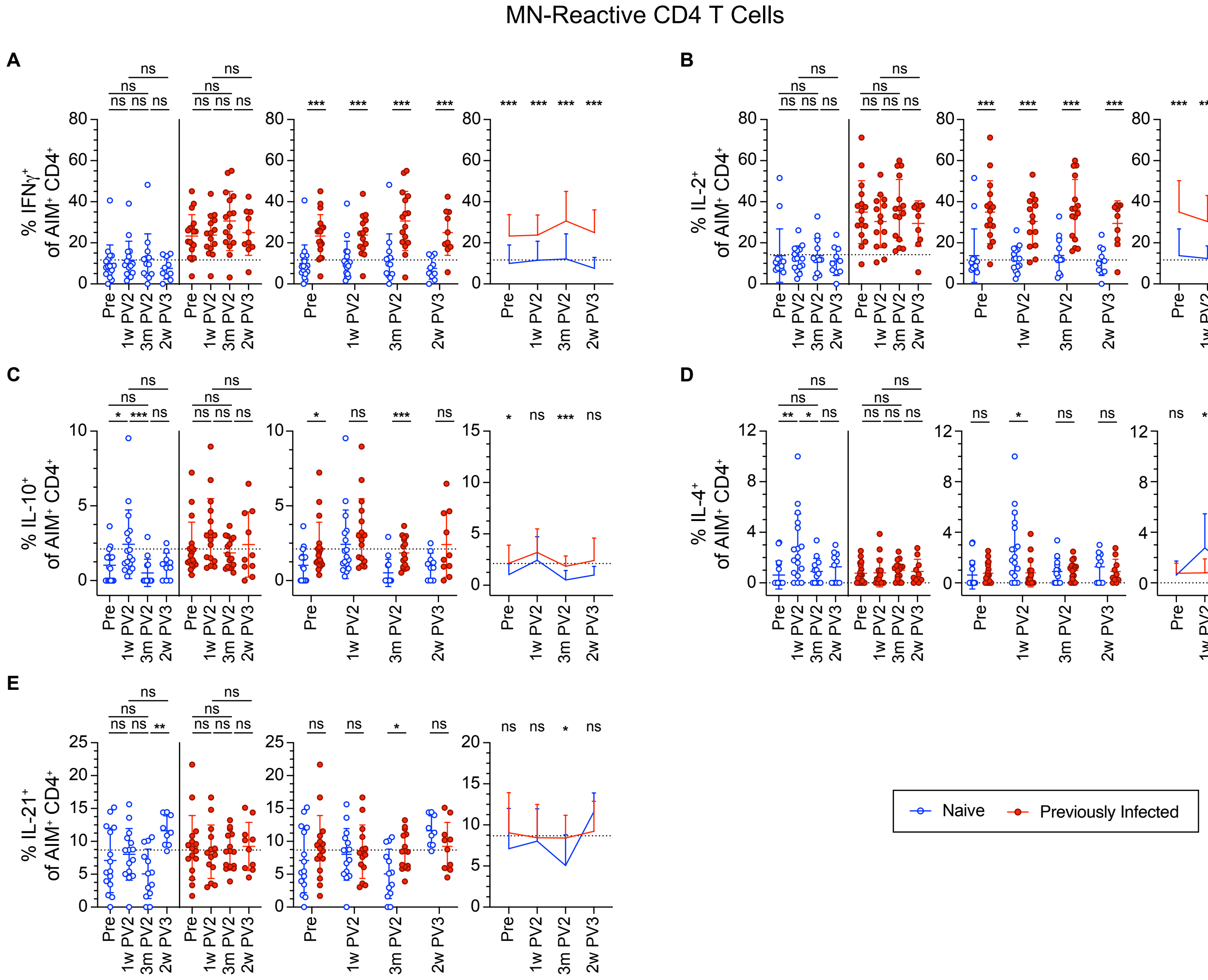

MN-Reactive CD4 T Cells
A
% IFNγ+ of AIM+ CD4+
B
% IL-2+ of AIM+ CD4+
C
% IL-10+ of AIM+ CD4+
D
% IL-4+ of AIM+ CD4+
E
% IL-21+ of AIM+ CD4+
Pre
1w PV2
3m PV2
2w PV3
ns
*
**
***
Naive
Previously Infected

Figure S11

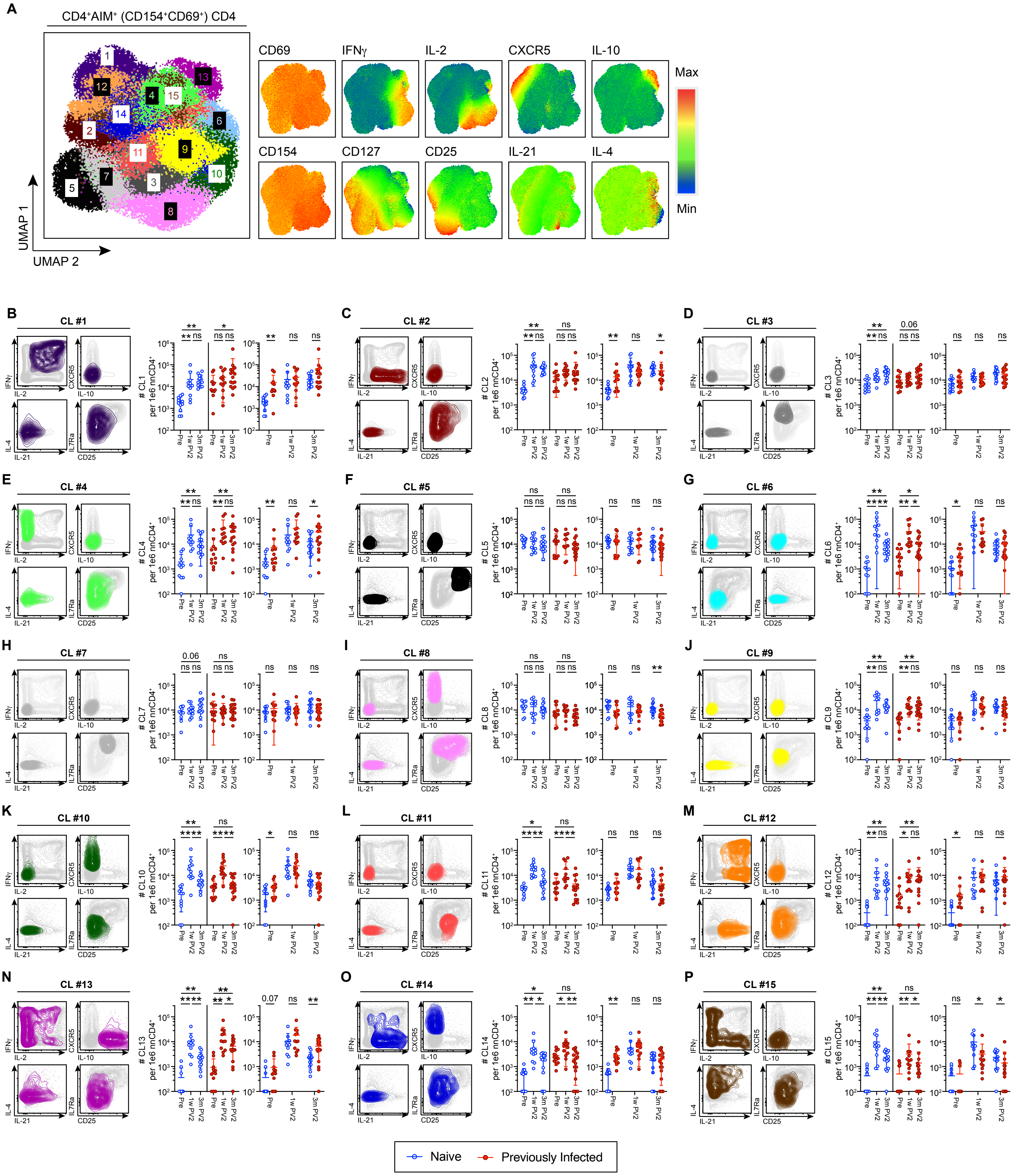
A
CD4+AIM+ (CD154+CD69+) CD4
UMAP 1
UMAP 2
CD69
IFNγ
IL-2
CXCR5
IL-10
CD154
CD127
CD25
IL-21
IL-4
Max
Min
B CL #1
C CL #2
D CL #3
E CL #4
F CL #5
G CL #6
H CL #7
I CL #8
J CL #9
K CL #10
L CL #11
M CL #12
N CL #13
O CL #14
P CL #15
per 1e6 nnCD4+
Pre
1w PV2
3m PV2
Naive
Previously Infected

#### Supplementary Table 1: Study cohort characteristics

| | **SARS-CoV-2 Naïve** | **SARS-CoV-2 Previously Infected** |
|---|---|---|
| Number of participants | 22 | 30 |
| Age (years) | 46 (25 – 79) | 50 (22 – 77) |
| Sex | 64% Female, 36% Male | 77% Female, 23% Male |
| Sustained symptoms (PASC)[1] | NA | 10 |
| Symptom onset to pre-vaccination draw (days) | NA | 265 (59 – 374) |
| Symptom onset to vaccine dose 1 (days) | NA | 305 (85 – 423) |
| Vaccine type[2] | 8 Moderna, 14 Pfizer | 10 Moderna, 20 Pfizer |
| Vaccine dose 2 to 1 week PV2 draw (days) | 9 (5 – 12) | 10 (5 – 20) |
| Vaccine dose 2 to 3 month PV2 draw (days) | 93 (75 – 119) | 91 (63 – 132) |
| Vaccine dose 2 to 6 month PV2 draw (days) | 161 (148 – 179) | 160 (133 – 190) |
| Vaccine dose 2 to vaccine dose 3 | 232 (182 – 274) | 233 (197 – 261) |
| Vaccine dose 3 to 2 week PV3 draw (days) | 19 (13 – 47) | 15 (10 – 22) |

[1] Participants surveyed at 6 months post-symptom onset for ongoing symptoms or post-acute sequelae of COVID-19 (PASC).

[2] mRNA-1273 (Moderna) or BNT162b2 (Pfizer-BioNTech)

### Supplementary Table 2 - T cell peptide pools

All Peptides from BEI product numbers NR-52402 (Spike), NR-52403 (Membrane), and NR-52404 (Nucleocapsid)

***Spike Peptide Pool***

| BEI Peptide number | Amino Acids | Sequence | HLA Class II epitope(s) | HLA Class I epitope(s) | AA sequence changes in variants |
|---|---|---|---|---|---|
| S-4 | 22-34 | TQLPPAYTNSFTRGVYY | | YTNSFTRGVY, LPPAYTNSF | |
| S-5 | 29-45 | TNSFTRGVYYPDKVFRS | | GVYYPDKVFR | |
| S-6 | 36-52 | VYYPDKVFRSSVLHSTQ | | KVFRSSVLH | |
| S-12 | 78-94 | RFDNPVLPFNDGVYFAS | | LPFNDGVYF | |
| S-16 | 106-122 | FGTTLDSKTQSLLIVNN | | TLDSKTQSL | |
| S-19 | 127-143 | VIKVCEFQFCNDPFLGV | CEFQFCNDPFLGVYY | | Οdel142-144, ΟY145D |
| S-20 | 134-150 | QFCNDPFLGVYYHKNNK | CEFQFCNDPFLGVYY | GVYYHKNNK | ΔG142D |
| S-23 | 155-171 | SEFRVYSSANNCTFEYV | SSANNCTFEYVSQPF | | ΔE156-, ΔF157-, ΔR158G |
| S-24 | 162-178 | SANNCTFEYVSQPFLMD | SSANNCTFEYVSQPF, CTFEYVSQPFLMDLE | | |
| S-25 | 169-185 | EYVSQPFLMDLEGKQGN | CTFEYVSQPFLMDLE | | |
| S-26 | 176-192 | LMDLEGKQGNFKNLREF | LMDLEGKQGNFKNLR | | |
| S-28 | 190-206 | REFVFKNIDGYFKIYSK | NIDGYFKIYSKHTPI | | |
| S-29 | 197-213 | IDGYFKIYSKHTPINLV | NIDGYFKIYSKHTPI | | Οdel211, ΟL212I |
| S-30 | 204-220 | YSKHTPINLVRDLPQGF | | TPINLVRDL | Οdel211, ΟL212I, Οins214EPE |
| S-31 | 211-227 | NLVRDLPQGFSALEPLV | NLVRDLPQGFSALEP | LPQGFSAL | ΔA222V, Οdel211, ΟL212I, Οins214EPE |
| S-33 | 225-241 | PLVDLPIGINITRFQTL | | LPIGINITRF | |
| S-39 | 267-283 | VGYLQPRTFLLKYNENG | | YLQPRTFLL | |
| S-42 | 288-304 | AVDCALDPLSETKCTLK | | ALDPLSETK | |
| S-46 | 316-332 | SNFRVQPTESIVRFPNI | | QPTESIVRF | |
| S-50 | 334-360 | ATRFASVYAWNRKRISN | RFASVYAWNRKRISN | SVYAWNRKR | ΟG339D |
| S-54 | 372-388 | ASFSTFKCYGVSPTKLN | | KCYGVSPTK | ΟS373P, ΟS375F |
| S-65 | 449-465 | YNYLYRLFRKSNLKPFE | | RLFRKSNLK | ΔL452R |
| S-66 | 456-472 | FRKSNLKPFERDISTEI | | KPFERDISTEI | |
| S-73 | 505-521 | YQPYRVVVLSFELLHAP | | QPYRVVVL, QPYRVVVLSF | ΟY505H |
| S-84 | 582-598 | LEILDITPCSFGGVSVI | | TPCSFGGVSV | ΔD614G |
| S-88 | 610-626 | VLYQDVNCTEVPVAIHA | | YQDVNCTEV | ΟD614G |
| S-99 | 687-703 | VASQSIIAYTMSLGAEN | | SIIAYTMSL | |
| S-102 | 708-724 | SNNSIAIPTNFTISVTT | | IPTNFTISV | |
| S-104 | 722-739 | VTTEILPVSMTKTSVDC | | EILPVSMTK | |
| S-108 | 750-766 | SNLLLQYGSFCTQLNRA | NLLLQYGSFCTQLNR | | ΟN764K |
| S-115 | 799-815 | GFNFSQILPDPSKPSKR | NFSQILPDPSKPSKR | | |
| S-119 | 827-843 | TLADAGFIKQYGDCLGD | AGFIKQYGDCLGDIA | TLADAGFIK | |
| S-120 | 834-850 | IKQYGDCLGDIAARDLI | AGFIKQYGDCLGDIA | | |
| S-123 | 855-871 | FNGLTVLPPLLTDEMIA | | GLTVLPPLL | ΟN856K |
| S-143 | 995-1011 | RLITGRLQSLQTYVTQQ | | RLQSLQTYV | |
| S-146 | 1016-1032 | AEIRASANLAATKMSEC | | RASANLAATK | |
| S-150 | 1044-1060 | GKGYHLMSFPQSAPHGV | | HLMSFPQSA, FPQSAPHGV | |
| S-152 | 1058-1074 | HGVVFLHVTYVPAQEKN | | VVFLHVTYV | |
| S-153 | 1065-1081 | VTYVPAQEKNFTTAPAI | | VTYVPAQEK | |
| S-163 | 1135-1151 | NTVYDPLQPELDSFKEE | | TVYDPLQPELDSFK | |
| S-180 | 1254-1270 | CKFDEDDSEPVLKGVKL | | EPVLKGVKL | |

***Membrane/Nucleocapsid Peptide Pool***

| BEI Peptide number | Amino Acids | Sequence | HLA Class II epitope(s) | HLA Class I epitope(s) | AA sequence changes in variants |
|---|---|---|---|---|---|
| M-3 | 15-31 | KLLEQWNLVIGFLFLTW | | KLLEQWNLV | OQ19E |
| M-6 | 36-52 | QFAYANRNRFLYIIKLI | QFAYANRNRFLYIIK | YANRNRFLY | |
| M-9 | 57-73 | LWPVTLACFVLAAVYRI | | FVLAAVYRI | |
| M-10 | 64-80 | CFVLAAVYRINWITGGI | VLAAVYRINWITGGI | | |
| M-11 | 71-87 | YRINWITGGIAIAMACL | YRINWITGGIAIAMA | | ΔI82T |
| M-13 | 85-101 | ACLVGLMWLSYFIASFR | CLVGLMWLSYFIASF, MWLSYFIASFRLFAR | | |
| M-14 | 92-108 | WLSYFIASFRLFARTRS | MWLSYFIASFRLFAR | | |
| M-16 | 106-122 | TRSMWSFNPETNILLNV | | SMWSFNPET | |
| M-17 | 113-129 | NPETNILLNVPLHGTIL | TNILLNVPLHGTILT | VPLHGTIL | |
| M-18 | 120-136 | LNVPLHGTILTRPLLES | TNILLNVPLHGTILT | | |
| M-19 | 127-143 | TILTRPLLESELVIGAV | | RPLLESEL | |
| M-20 | 134-150 | LESELVIGAVILRGHLR | SELVIGAVILRGHLR | | |
| M-21 | 141-157 | GAVILRGHLRIAGHHLG | RGHLRIAGHHLGRCD | | |
| M-22 | 148-164 | HLRIAGHHLGRCDIKDL | RGHLRIAGHHLGRCD, IAGHHLGRCDIKDLP | RIAGHHLGR | |
| M-23 | 155-171 | HLGRCDIKDLPKEITVA | AGHHLGRCDIKDLP, LGRCDIKDLPKEITV, IKDLPKEITVATSRT | | |
| M-24 | 162-178 | KDLPKEITVATSRTLSY | IKDLPKEITVATSRT, KEITVATSRTLSYYK | LPKEITVAT | |
| M-25 | 169-185 | TVATSRTLSYYKLGASQ | KEITVATSRTLSYYK | | |
| M-26 | 176-192 | LSYYKLGASQRVAGDSG | LSYYKLGASQRVAGD | | |
| M-27 | 183-199 | ASQRVAGDSGFAAYSRY | | AGDSGFAAY, RVAGDSGFAAY | |
| M-28 | 190-206 | DSGFAAYSRYRIGNYKL | SGFAAYSRYRIGNYK | | |
| N-1 | Start-17 | MSDNGPQNQRNAPRITF | | GPQNQRNAPRITF | OP13L |
| N-6 | 36-52 | RSKQRRPQGLPNNTASW | | RPQGLPNNTA | |
| N-7 | 43-59 | QGLPNNTASWFTALTQH | | LPNNTASWF | |
| N-8 | 50-66 | ASWFTALTQHGKEDLKF | SWFTALTQHGKEDLK | | ΔD63G |
| N-10 | 64-80 | LKFPRGQGVPINTNSSP | | FPRGQGVPI | |
| N-11 | 71-87 | GVPINTNSSPDDQIGYY | | NTNSSPDDQIGYY | |
| N-12 | 78-94 | SSPDDQIGYYRRATRRI | DDQIGYYRRATRRIR | | |
| N-13 | 85-101 | GYYRRATRRIRGGDGKM | DDQIGYYRRATRRIR, YYRRATRRIRGGDGK | | |
| N-15 | 99-115 | GKMKDLSPRWYFYYLGT | | SPRWYFYYL | |
| N-18 | 120-136 | GLPYGANKDGIIWVATE | NKDGIIWVATEGALN | | |
| N-19 | 127-143 | KDGIIWVATEGALNTPK | NKDGIIWVATEGALN | | |
| N-22 | 148-164 | TRNPANNAAIVLQLPQG | | NPANNAAIVL | |
| N-31 | 211-227 | AGNGGDAALALLLLDRL | AGNGGDAALALLLLD, DAALALLLLDRLNQL | | |
| N-32 | 218-234 | ALALLLLDRLNQLESKM | DAALALLLLDRLNQL, LLLLDRLNQLESKMS | LLLDRLNQL | |
| N-33 | 225-241 | DRLNQLESKMSGKGQQQ | LLLLDRLNQLESKMS | | |
| N-36 | 246-262 | VTKKSAAEASKKPRQKR | AAEASKKPRQKRTAT | | |
| N-37 | 253-269 | EASKKPRQKRTATKAYN | AAEASKKPRQKRTAT, KKPRQKRTATKAYNV | KPRQKRTAT | |
| N-38 | 260-276 | QKRTATKAYNVTQAFGR | KKPRQKRTATKAYNV, KRTATKAYNVTQAFG | | |
| N-43 | 295-311 | GTDYKHWPQIAQFAPSA | WPQIAQFAPSASAFF | | |
| N-44 | 302-318 | PQIAQFAPSASAFFGMS | WPQIAQFAPSASAFF | APSASAFFGM | |
| N-45 | 309-325 | PSASAFFGMSRIGMEVT | | GMSRIGMEV | |
| N-47 | 316-332 | GMSRIGMEVTPSGTWLT | PSGTWLTYTGAIKLD | | |
| N-48 | 323-339 | EVTPSGTWLTYTGAIKL | PSGTWLTYTGAIKLD | | |
| N-50 | 344-360 | PNFKDQVILLNKHIDAY | FKDQVILLNKHIDAY | | |
| N-51 | 351-367 | ILLNKHIDAYKTFPPTE | ILLNKHIDAYKTFPPTE | | |
| N-52 | 358-374 | DAYKTFPPTEPKKDKKK | | KTFPPTEPKK, KTFPPTEPKKDKKK | |

#### Supplementary Table 3 - Key Resources

| Reagent or Resource | Source | Identifier |
|---|---|---|
| *Antibodies* | | |
| Anti-human CD45-BUV395, Clone HI30 | Becton Dickinson | 563792 |
| Anti-human CD45-BUV496, Clone HI30 | Becton Dickinson | 750179 |
| Anti-human CD3-BUV615, Clone UCHT1 | Becton Dickinson | 612993 |
| Anti-human HLA-DR-BUV661, Clone G46-6 | Becton Dickinson | 612981 |
| Anti-human CD45RA-BUV737, Clone HI100 | Becton Dickinson | 612847 |
| Anti-human CD26-BUV805, Clone M-A261 | Becton Dickinson | 749316 |
| Anti-human CXCR3-BV421, Clone G025H7 | Biolegend | 353716 |
| Anti-human CD45-e450, Clone HI30 | Thermo Fisher | 49-0459-42 |
| Anti-human CD8a-BV480, Clone RPA-T8 | Becton Dickinson | 566163 |
| Anti-human CCR7-BV605, Clone G043H7 | Biolegend | 353224 |
| Anti-human CCR6-BV650, Clone G043G3 | Biolegend | 353426 |
| Anti-human CD27-BV711, Clone M-T271 | Biolegend | 356430 |
| Anti-human CD137-BV750, Clone 4B4-1 | Biolegend | 309844 |
| Anti-human CD57-BV785, Clone QA17A04 | Biolegend | 393329 |
| Anti-human CXCR5-BB515, Clone RF8B2 | Becton Dickinson | 564624 |
| Anti-human CD45-Ax532, Clone HI30 | Thermo Fisher | 58-0459-42 |
| Anti-human CD134-PerCP/cy5.5, Clone BerACT35 | Biolegend | 350010 |
| Anti-human PD-L1-PE, Clone 29E.2A3 | Biolegend | 329705 |
| Anti-human CCR4-PE/Dazzle594, CLone L291H4 | Biolegend | 359420 |
| Anti-human CD25-PE/Cy5, Clone BC96 | Biolegend | 302608 |
| Anti-human CD127-PE/Cy7, Clone hIL7Rm21 | Biolegend | 560822 |
| Anti-human ICOS-APC, Clone C398.4a | Biolegend | 313509 |
| Anti-human CD4-SparkNIR685, Clone SK3 | Biolegend | 344657 |
| Anti-human CD69-APC/R700, Clone FN50 | Becton Dickinson | 565154 |
| Anti-human gdTCR-APC/Fire750, Clone B1 | Biolegend | 331228 |
| Anti-human CD19-APC/Fire810, Clone HIB19 | Biolegend | 302271 |
| Anti-human CD69-BUV395, Clone FN50 | Becton Dickinson | 564364 |
| Anti-human IL-13 BV421, Clone JES10-5E2 | Biolegend | 501916 |
| Anti-human CD3 eF450, Clone OKT3 | eBioscience | 48-0037-42 |
| Anti-human CD107a BV510, Clone H4A3 | Biolegend | 328632 |
| Anti-human IL-17A BV570, Clone BL168 | Biolegend | 512324 |
| Anti-human CD154 Biotin, Clone hCD40L-M91 | Becton Dickinson | 552560 |
| Streptavidin BV605, Clone 563260 | Becton Dickinson | 563260 |
| Anti-human CD25 BV650, Clone M-A251 | Becton Dickinson | 563719 |
| Anti-human CD45RA BV711, Clone HI100 | Becton Dickinson | 563733 |
| Anti-human CD19 BV711, Clone SJ25C1 | Becton Dickinson | 563036 |
| Anti-human CD16 BV711, Clone 3G8 | Becton Dickinson | 563127 |
| Anti-human CD14 BV711, Clone M5E2 | Biolegend | 301838 |
| Anti-human IL-2 BV785, Clone MQ1-17H12 | Biolegend | 500348 |
| Anti-human CD127 AlexaFlour 488, Clone AO19D5 | Biolegend | 351314 |
| Anti-human IL-21 PE, Clone 3A3-N2 | eBioscience | 12-7219-42 |
| Anti-human IL-10 PE Dazzle 594, Clone JES3-9D7 | Biolegend | 501426 |
| Anti-human CD8 PECy5, Clone RPA-T8 | Becton Dickinson | 555368 |
| Anti-human IL-4 PECy7, Clone MP4-25D2 | Biolegend | 500824 |
| Anti-human CXCR5 Alexa Flour 647, Clone J252D4 | Biolegend | 356906 |
| Anti-human CD4 Alexa Flour 700, Clone RPA-T4 | Becton Dickinson | 557922 |

| | | |
|---|---|---|
| Anti-human IFNy APCeF780, Clone 4s-B3 | eBioscience | 47-7319-42 |
| Anti-human CD3-PerCpCy5.5, Clone HIT3a | BioLegend | 300328 |
| Anti-human CD14-PerCpCy5.5, Clone M5E2 | BioLegend | 301824 |
| Anti-human CD16-PerCpCy5.5, Clone 3G8 | BioLegend | 302028 |
| Anti-human CD19-BUV496, Clone SJ25C1 | BD | 612939 |
| Anti-human CD20-BV711, Clone 2H7 | BioLegend | 302341 |
| Anti-human IgM-BV510, Clone MHM-88 | BioLegend | Cat #314521 |
| Anti-human IgD-BUV395, Clone IA6-2 | BD | 563813 |
| Anti-human IgG-BV786, Clone G18-145 | BD | Cat #564230 |
| Anti-human CD21-SB600 Clone, HB5 | ThermoFisher | Cat #63-0219-41 |
| Anti-human CD27-BV421, Clone M-T271 | BioLegend | Cat #356418 |
| Anti-human CD38-AF700, Clone HIT2 | BD | Cat #56-0381-82 |
| Anti-human IgA-Biotin, Clone IS11-8E10 | Miltenyi | Cat #130-113-474 |
| Anti-human IgA-PE-Vio 770, Clone IS11-8E10 | Miltenyi | Cat #130-114-003 |
| Anti-human CD11c-PEDazzle594, Clone 3.9 | BioLegend | Cat #301642 |
| Anti-human IgG-HRP | Jackson ImmunoResearch | Cat #109-035-088 |
| anti-human IgA-HRP | Southern Biotech | Cat #2050-05 |
| *Bacterial and Virus Strains* | | |
| SARS-CoV-2 (WA-1) | BEI resources | NR-52281 |
| *Biological Samples* | | |
| Human PBMC | This paper | N/A |
| *Chemicals, Peptides, and Recombinant Proteins* | | |
| Zombie LiveDead NIR | Biolegend | 423105 |
| Fixable Live Dead BLUE BUV496 | Thermo Scientific | |
| Decoy Tetramer PE/Dy594/Dy650 | This paper | N/A |
| SA-PE | Agilent | Cat#PJRS301-1 |
| RBD Tetramer PE | This paper | N/A |
| BirA500 kit | Avidity | Cat #BirA500 |
| anti-PE magnetic beads | Miltenyi Biotec | Cat#130-048-801 |
| Streptavidin-BUV805 | BD | 564923 |
| phorbol 12-myristate 13-acetate | Sigma-Aldrich | Cat#P8139 |
| Ionomycin | Sigma-Aldrich | Cat#I9657 |
| GolgiStop/monensin | Becton Dickinson | Cat#554724 |
| Paraformaldahyde solution, 4% in PBS | Thermo Scientific | Cat#AAJ19943K2 |
| Cytofix/Cytoperm | Becton Dickinson | Cat#554714 |
| DMSO, Cell culture grade >99.5% | Sigma-Aldrich | Cat#D4540 |
| CEFX Ultra SuperStim Pool | JPT | Cat#PM-CEFX-2 |
| SARS-CoV-2 HLA Class I & II 15-mer peptides: Membrane | BEI Resources | Cat#NR-52403 |
| SARS-CoV-2 HLA Class I & II 15-mer peptides: Nucleocapsid | BEI Resources | Cat#NR-52404 |
| SARS-CoV-2 HLA Class I & II 15-mer peptides: Spike | BEI Resources | Cat#NR-52402 |
| Dylight NHS Ester 594 | ThermoFisher | 46413 |
| Dylight NHS Ester 650 | ThermoFisher | 62266 |
| 1X 3,3′,5,5′-Tetramethylbenzidine (TMB) | Invitrogen | Cat#00-4201-56 |
| Recombinant SARS-CoV-2 (Wuhan-1) RBD Protein | Walls et al, 2009 | N/A |
| anti-CD40 agonist mAb | Miltenyi | 130-094-133 |
| *Software and Algorithms* | | |

| | | |
|---|---|---|
| FlowJo10 | Becton Dickinson | N/A |
| SpectroFlo | Cytek Biosciences | N/A |
| Prism | GraphPad | N/A |
| R | R core team | N/A |
